# Ensemble Learning: Predicting Human Pathogenicity of Hematophagous Arthropod Vector-Borne Viruses

**DOI:** 10.1101/2023.12.30.23300660

**Authors:** Huakai Hu, Chaoying Zhao, Meiling Jin, Jiali Chen, Xiong Liu, Hua Shi, Jinpeng Guo, Changjun Wang, Yong Chen

**Affiliations:** School of Public Health, China Medical University, Shenyang, Liaoning province, China; China Chinese PLA Center for Disease Control and Prevention, Beijing, China; School of Public Health, Zhengzhou University, Zhengzhou, Henan Province, China; Liaoning Provincial Center for Disease Control and Prevention, Shenyang, Liaoning, People’s Republic of China; School of Medicine, NanKai University, Tianjin, People’s Republic of China

## Abstract

Hematophagous arthropods serve as crucial vectors for numerous viruses, posing significant public health risks due to their potential for zoonotic spillover. Despite the advances in metagenomics expanding our understanding of arbovirus diversity, traditional phylogenetic approaches often miss the pathogenic potential of viruses not yet identified in humans. Here, we curated two datasets: one with 294 viruses and 36 epidemiological characteristics (including virus properties, vector hosts, and non-vector hosts), and another with 71,622 viral sequences focusing on pathogenic traits. Using these datasets, we developed a regression model and a prediction model to assess and predict viral pathogenicity. Using these datasets, we developed a regression model and a prediction model to assess and predict viral pathogenicity. Our regression model (R^2^ = 90.6%) reveals a strong correlation between non-vector host diversity, especially within *Perissodactyla* and *Carnivora* orders, and virus pathogenicity. The prediction model (F1 score = 96.79%) identifies key pathogenic functions such as “Viral adhesion” and “Host xenophagy” as enhancers of pathogenic potential, while the “Viral invasion” function was associated with an inverse effect. Validation against an external independent dataset confirmed the models’ ability to identify pathogenic viruses and revealed the potential threat posed by Palma and Zaliv Terpeniya viruses, previously undetected in humans. These findings highlight the necessity of integrating predictive models with metagenomic data to provide early warnings of potential zoonotic viruses carried by hematophagous vectors at the strain level, enhancing public health responses and preparedness.

## Introduction

Hematophagous arthropods, such as mosquitoes and ticks, play a crucial role in ecosystems as blood consumers and disease vectors (Cuthbert et al., 2023; Touray et al., 2023). They can harbor various pathogens, including bacteria, fungi, and viruses. Viral infections in these organisms are classified as Arthropod-Borne Viruses (arboviruses) and insect-specific viruses (ISVs) (Calisher & Higgs, 2018; Gould et al., 2017; Nouri et al., 2018; Zhao et al., 2022). Arthropods have the ability to carry and spread various pathogens, which pose a significant threat to the health of humans and animals. This can lead to outbreaks and an increase in annual mortality rates (Batson et al., 2021; Chala & Hamde, 2021; Roth et al., 2018). Notable arboviruses include Zika virus (ZIKAV) (Khongwichit et al., 2023; Weaver et al., 2018), Japanese encephalitis virus (JEV) (Kampen & Werner, 2014), and the incessant menace of Dengue virus (DENV) (Fournet et al., 2023). In recent years, propelled by the widespread adoption of viral metagenomics sequencing technologies, the identification of a wide range of established and emerging viruses within hematophagous vectors, such as mosquitoes and ticks, has become feasible (Ni et al., 2023; X. Yang et al., 2023). This technological progress presents an unprecedented opportunity to comprehensively explore the distribution and transmission patterns of arboviruses and ISVs across a spectrum of hosts, including both vectors and non-vectors. Such advancements are crucial for supporting early warning systems, facilitating the anticipation and mitigation of disease spread before its onset (Birnberg et al., 2020; Brinkmann et al., 2016). Despite these achievements in viral metagenomics, current bioinformatic methods for virus recognition still face limitations (Fang et al., 2019). Accurate identification of a significant number of unknown contigs remains challenging. Particularly when identifying known or novel viruses, the direct isolation and cultivation of these viruses from vectors proves to be formidable tasks, hindering in-depth exploration of their pathogenesis and immune response (Lewis et al., 2021).

In general, the close phylogenetic relatedness among viruses can provide insights into their potential for human infectivity, as closely related viruses are generally presumed to share common phenotypes and host ranges (Geoghegan & Holmes, 2018). However, despite being a common rule of thumb for virus risk assessment, the extent to which evolutionary proximity to viruses with known human infectivity accurately predicts zoonotic potential remains unexamined in the current literature (Behl et al., 2022). A model is specifically designed to utilize sequence features of closely related viruses (i.e., strains of the same species) to discern their potential for human infectivity (Zhang et al., 2019). However, this approach may overlook critical functional information of the viral genome, leading to a model that might not reliably identify pathogenic features applicable across diverse viruses. Consequently, pathogenicity predictions derived from such a model could be prone to significant biases (Mollentze et al., 2021).

The transmission of a virus is influenced by various epidemiological characteristics, including not only the virus itself and its vector host (Zaid et al., 2021; Y.-J. S. Huang et al., 2019a; Viglietta et al., 2021), but also geographical and climatic conditions, as well as interactions with non-vector hosts (Ciota & Keyel, 2019; Conway et al., 2014; Forrester et al., 2014; Tabachnick, 2016). The nucleotide sequence information of specific viruses holds importance, as it can provide insights into the underlying pathogenic mechanisms (Bartoszewicz, Genske, et al., 2021). By identifying these pathogenic viruses early, we are enabled to establish more effective monitoring and early warning systems. Drawing on the global dataset of arboviruses and ISVs compiled by Huang et al. (Y. Huang et al., 2023), we curated a dataset encompassing viruses transmitted by hematophagous arthropods, along with their associated epidemiological data. In parallel, we created another dataset by utilizing the pathogenic functionalities within viral sequences discerned by SeqScreen (Balaji et al., 2022). Employing these two datasets and leveraging the XGBoost algorithm within an ensemble learning framework, regression and classification models were developed. Our aim was to identify the human pathogenicity of arboviruses and evaluate the zoonotic spillover risk for earlier public health prevention and control.

## Materials and methods

### Database restructuring and epidemiological feature retrieval

The database initially comprised 101,094 virus sequences sourced from NCBI and GenBank, covering the period from March 11, 1991, to January 28, 2023 (Y. Huang et al., 2023). We then compiled a table of 11 species of hematophagous arthropods, including mosquitoes, ticks, and sand flies, while excluding non-blood-feeding species such as *Tipulidae* and *Chironomidae* (Supplementary Table 1). To enhance precision and specificity, a stringent screening process was applied, systematically excluding records that lacking host information, sampling location details, as well as those with ambiguous vector-host relationships or vectors originating from Antarctica. Following this screening process, we obtained a database focused on viruses transmitted by hematophagous arthropods. Additionally, we developed a Python script to extract detailed taxonomic information for non-vector hosts at the order, family, and genus levels. To facilitate the discernment of relative differences among vector hosts across different countries, we performed a logarithmic transformation on the counts of hosts (Figure 1A). Moreover, to emphasize the primary non-vector hosts, those recorded fewer than 100 times were aggregated into an “Others” category, yielding a total of 10 classifications (Figure 1C).

**Figure 1:**
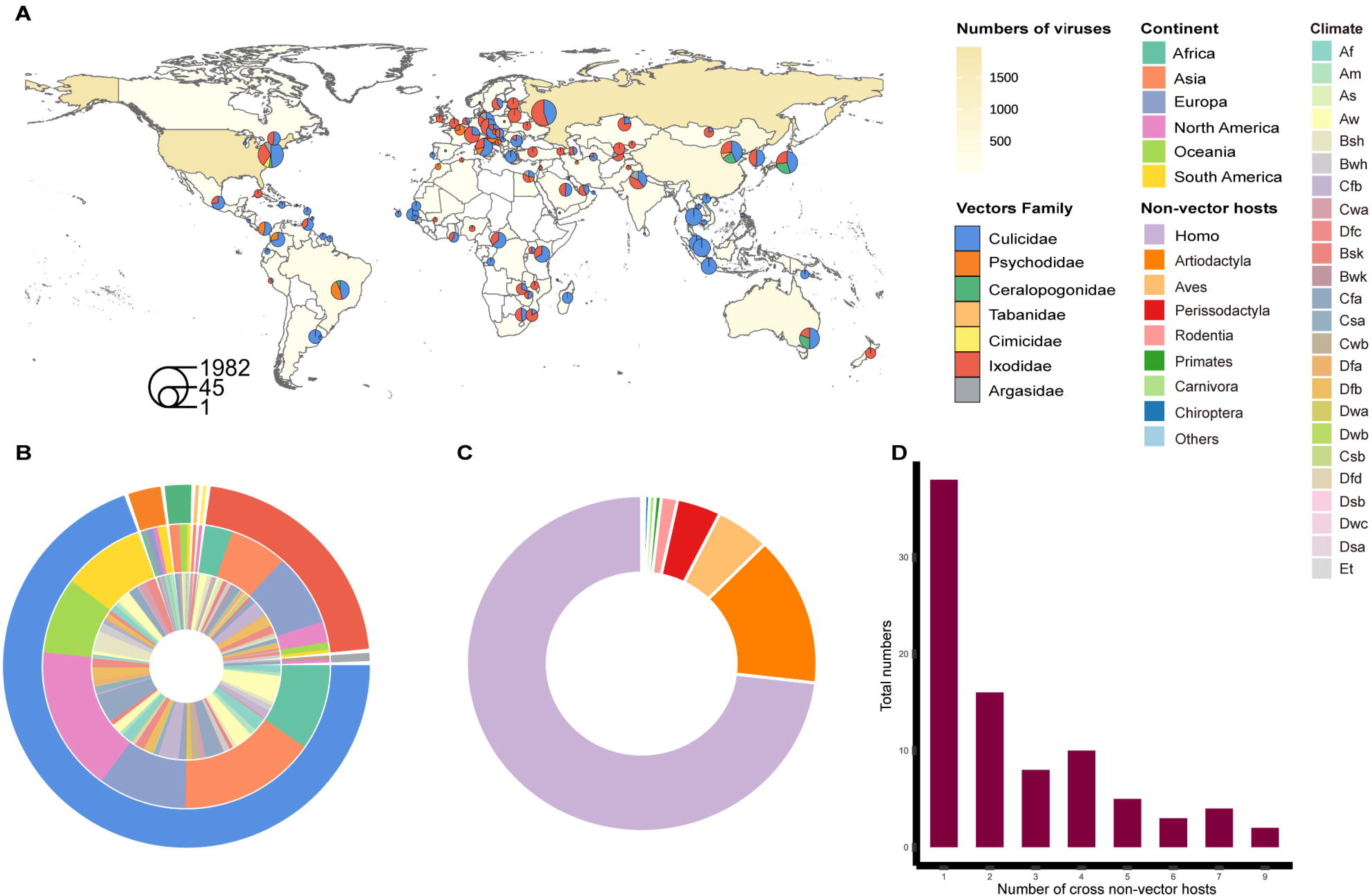
Global characteristics of hematophagous arthropod vectors and non-vector hosts for arboviruses and ISVs. (A) Global distribution and quantity of hematophagous arthropod vectors. (B) The diversity of vector hosts, their geographic locations across continents, and their prevalence in various climatic conditions. (C) Diversity and quantity of non-vector hosts associated with arboviruses and ISVs. (D) Distribution of viruses across non-vector hosts: the horizontal axis represents the number of non-vector hosts a virus can infect, and the vertical axis indicates the total number of such viruses.

Additional epidemiological features were added to the basic database to enhance our analysis. Specifically, the geographical location of each arthropod vector was acquired from the World Population Review (https://worldpopulationreview.com/continents) to refine our geographical analysis. Subsequently, the Köppen climate classification was determined for each vector based on its location of discovery, using data sourced from the Weather and Climate website (https://weatherandclimate.com/) and the Mindat website (https://www.mindat.org/). Furthermore, the Baltimore classification data for the viruses were integrated into our database, sourced from the International Committee on Taxonomy of Viruses (ICTV) (https://ictv.global/report/genome).

### The development of a regression model for epidemiological characteristics

After the screening process, we obtained a dataset of 8,366 records, each enriched with unique epidemiological characteristics. The dataset for the regression model was transformed using R, which included 294 viruses and 36 epidemiological characteristics (Supplementary Table 2). The XGBoost was used to develop regression models. The dataset was partitioned into training and validation sets at a ratio of 7.5:2.5. The model’s independent variables consist of 36 epidemiological characteristics, with the dependent variable representing a binary indicator of human infection status (0 = no, 1 = yes). Considering that positive samples represented 55% of the total, adjustments were made to maintain the balance of the dataset during construction. The model was trained on the training set using specified parameters (Supplementary Table 3) and the optimal number of iterations was determined through 10-fold cross-validation. Following this, the final model was constructed. The effectiveness of the model was assessed by means of the validation set, using Mean Absolute Error (MAE) and R^2^ as the main evaluation metrics.

### The development of a classification model for viral sequence pathogenic functionality

Pathogenic functionality information was obtained for 71,623 viral sequences through the annotation of the database using SeqScreen (Balaji et al., 2022). Among these, the pathogenicity function data of 228 viruses, which were carried by arthropod vectors and submitted to NCBI after 2022, were utilized as an external validation dataset for verification of the model’s prediction results. The remaining 71,395 viruses served as the training dataset for the classification model. Within the model dataset, data were allocated into training and testing sets at a 7.5:2.5 ratio. This model incorporates a total of 11 independent variables, including 10 pathogenic function features and sequence size. The dependent variable represents a binary indicator of human infection status (0 = no, 1 = yes). Owing to the disparity between positive and negative samples in the dataset (positivity rate of 79.68%), the sampling rate was adjusted to achieve balance. Subsequently, the model underwent training on the allocated training set using specified parameters (Supplementary Table 4), with the optimal number of iterations determined through 10-fold cross-validation. Furthermore, the Arbovirus Human Pathogen Prediction (AHPP) model was constructed utilizing the identified optimal iteration count. Precision and F1 score were used to evaluate the model’s performance.

To predict the human pathogenicity of viruses within an external validation dataset, two distinct prediction models were employed. The outcomes of these predictions were then compared and validated to assess the capabilities of our models. Among these, the zoonotic rank model is designed to evaluate the zoonotic risk associated with viruses (Mollentze et al., 2021). According to this model, ratings of “High” and “Very high” indicate a significant risk of zoonosis, suggesting that the virus poses a considerable threat of cross-species transmission to humans. Conversely, ratings of “Medium” and “Low” suggest a lower likelihood of zoonotic risk.

## Results

### Global distribution, diversity, and host interactions of hematophagous arthropod-borne viruses

This study curated a detailed dataset comprising 8,468 pairs of hematophagous vectors and viruses, elucidating their geographic distribution, diversity, and host interactions. In terms of distribution, the vectors fall into two principal classes: *Insecta* and *Arachnida*, across seven distinct families (Figure 1A). The dataset includes records from all six inhabited continents, with the notable exception of Antarctica, covering 102 countries globally and representing 24 diverse climate types (Figure 1B). Regarding the diversity among these vectors, the *Culicidae* family dominates with 5,445 sequences, representing 64% of the dataset. It is followed by the *Ixodidae* family, which comprises 2,703 sequences, representing 32%. The United States exhibits the greatest diversity and abundance of vectors, hosting representatives from five distinct families. China ranks second, with vectors from four families. Concerning virus records associated with vectors, the United States leads with 1,977 records, with Russia, China, and Japan sequentially trailing.

Concerning non-vector hosts, the dataset includes an additional 54,789 pairs of non-vector hosts and viruses, which fall into 15 distinct groups. Among these, humans are the most common, represented by 40,078 records, followed by *Artiodactyla* and *Aves*, which together account for nearly 20% of the total (Figure 1C). While most arboviruses and ISVs tend to associate with a singular non-vector host, exceptions such as the West Nile virus (WNV) and Tick-borne encephalitis virus (TBEV) from the *Flaviviridae* family demonstrate more extensive cross-host transmission, detected in nine non-vector host species. As the diversity of non-vector hosts a virus can infect increases, there is a notable trend toward a reduction in the number of virus species (Figure 1D). Specifically, viruses that infect only one host span ten different virus families, while those capable of infecting two to four hosts are limited to five families. Viruses with the capability to infect five to seven hosts are predominantly from the Flaviviridae and Togaviridae families. Furthermore, a marked correlation exists in the detection numbers among different viruses. This pattern is particularly evident among viruses from the *Togaviridae*, *Flaviviridae*, *Peribunyaviridae*, and *Phenuiviridae* families, which are often co-detected (Figure S1). Such findings highlight the complex interrelationships and dynamics within virus-vector host interactions.

### Human pathogenicity of hematophagous arthropod vector-borne viruses: a regression analysis of epidemiological characteristics

This section presents the dataset constructed for 294 viruses, which includes 36 epidemiological characteristics grouped into three categories: characteristics inherent to the virus, characteristics of vector hosts, and characteristics of non-vector hosts. The regression model developed herein exhibited strong performance on the test set, evidenced by a Mean Squared Error (MSE) of 2.3% and an R^2^ of 90.6%. These results suggest that the included variables are capable of significantly elucidating the factors contributing to the human pathogenicity of these viruses.

Our analysis reveals that the diversity of non-vector hosts for vector-borne viruses is significantly associated with increased human pathogenicity, surpassing the influence of both arthropod vector hosts and the viruses’ inherent properties (Figure 2). This characteristic is emphasized by its prominent position in both the model’s Gain and Cover metrics (Figure S2). *Perissodactyla* and *Carnivora*, as non-vector hosts, are particularly significant. The frequent detection of viruses in these species strongly correlates with a heightened risk of spillover events. Equally pivotal is the role of vector hosts. Viruses present in diverse vector genera are associated with heightened pathogenic risks. Ticks, for example, are known to transmit a significant number of harmful viruses. Furthermore, vector-borne viruses, especially those classified as arboviruses or within the Flaviviridae family, are significant in facilitating the transmission of diseases to humans.

**Figure 2:**
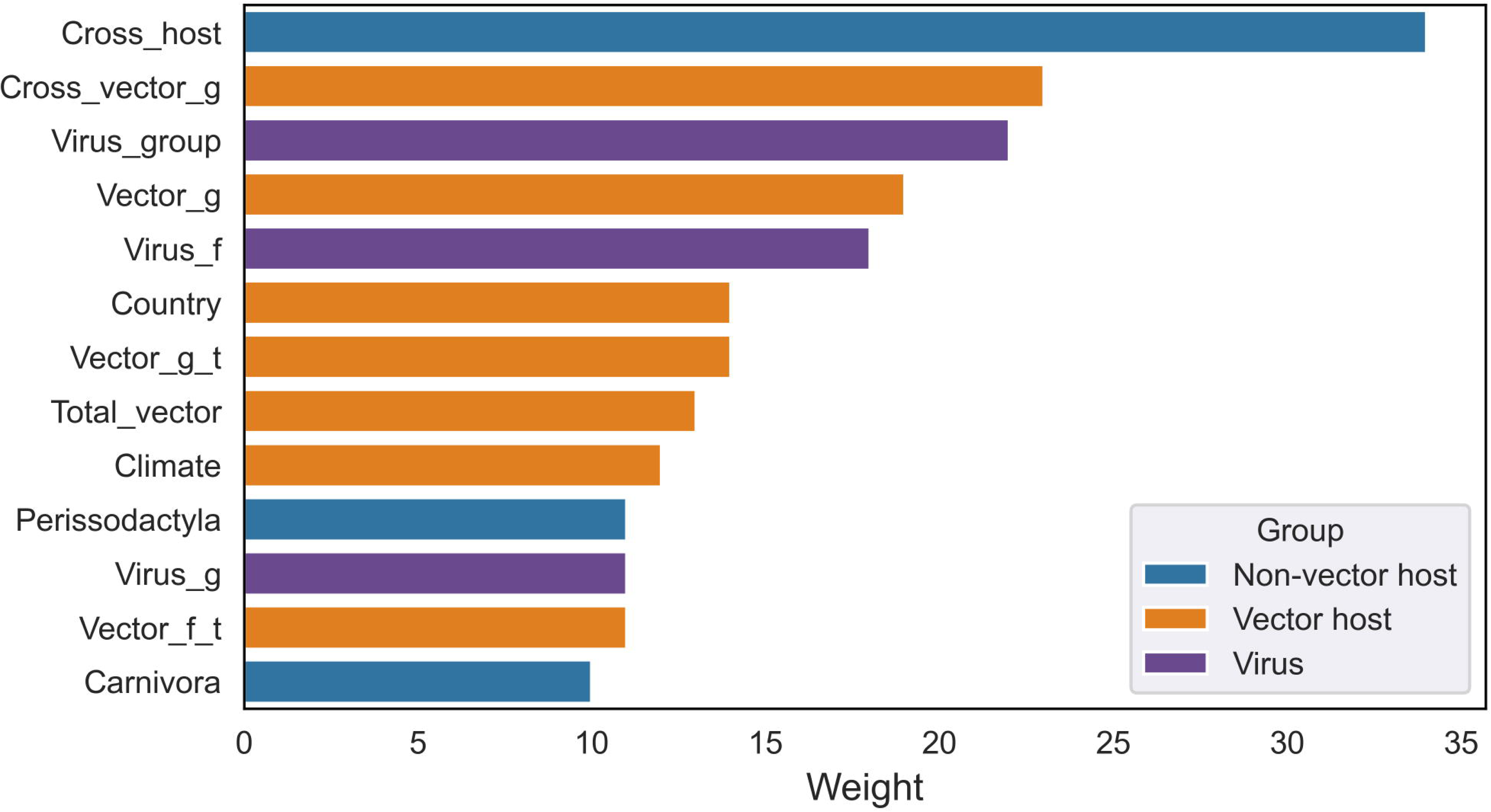
Importance of epidemiological characteristics for human pathogenicity of viruses. The weighted contributions of various epidemiological features to the prediction of human pathogenicity within the framework of an XGBoost regression model.

### Human Pathogenicity and Pathogenic Function of Virus Sequences: A Classification Approach

In our research, SeqScreen was employed to annotate the pathogenic functionality of all viral sequences within our database. Following the exclusion of sequences that failed to be successfully annotated, our dataset was refined to include a total of 71,623 sequences, encompassing both arboviruses and ISVs. Each characterized by pathogenic features, host information, and established zoonotic potential. Mosquito-borne arboviruses formed a significant portion of the dataset, with DENV-1 (9,194 sequences), DENV-2 (8,999 sequences), and West Nile virus (WNV) (4,656 sequences) from the *Flaviviridae* family being the most prevalent. Tick-borne arboviruses were also well-represented, including African swine fever virus (ASFV) (3,915 sequences) from the *Asfarviridae* family and Crimean-Congo hemorrhagic fever orthonairovirus (CCHF) (3,771 sequences) from the *Nairoviridae* family, also contributing a notable proportion to the dataset.

After screening an initial set of 32 features, ten distinct pathogenic features were identified within these sequences (Table 1). Among these, “Viral adhesion” emerged as the most prevalent, accounting for 62% (44,482 sequences) of the total. This function is crucial for the attachment of viruses to host cell surface receptors, marking the first step in the infection process that facilitates viral entry and sets the stage for further viral invasion and replication inside the cell. “Viral counter signaling” and “Host xenophagy” are also significant, accounting for 49% and 47% of the features, respectively, and are primarily associated with the virus’s immune evasion strategies. The former disrupts host immune signaling pathways to prevent inflammatory responses and other immune defenses, while the latter involves the virus’s interference with the host’s autophagic processes. Together, both features significantly prolong the virus’s survival time within host cells by protecting viral integrity.

**Table 1:**
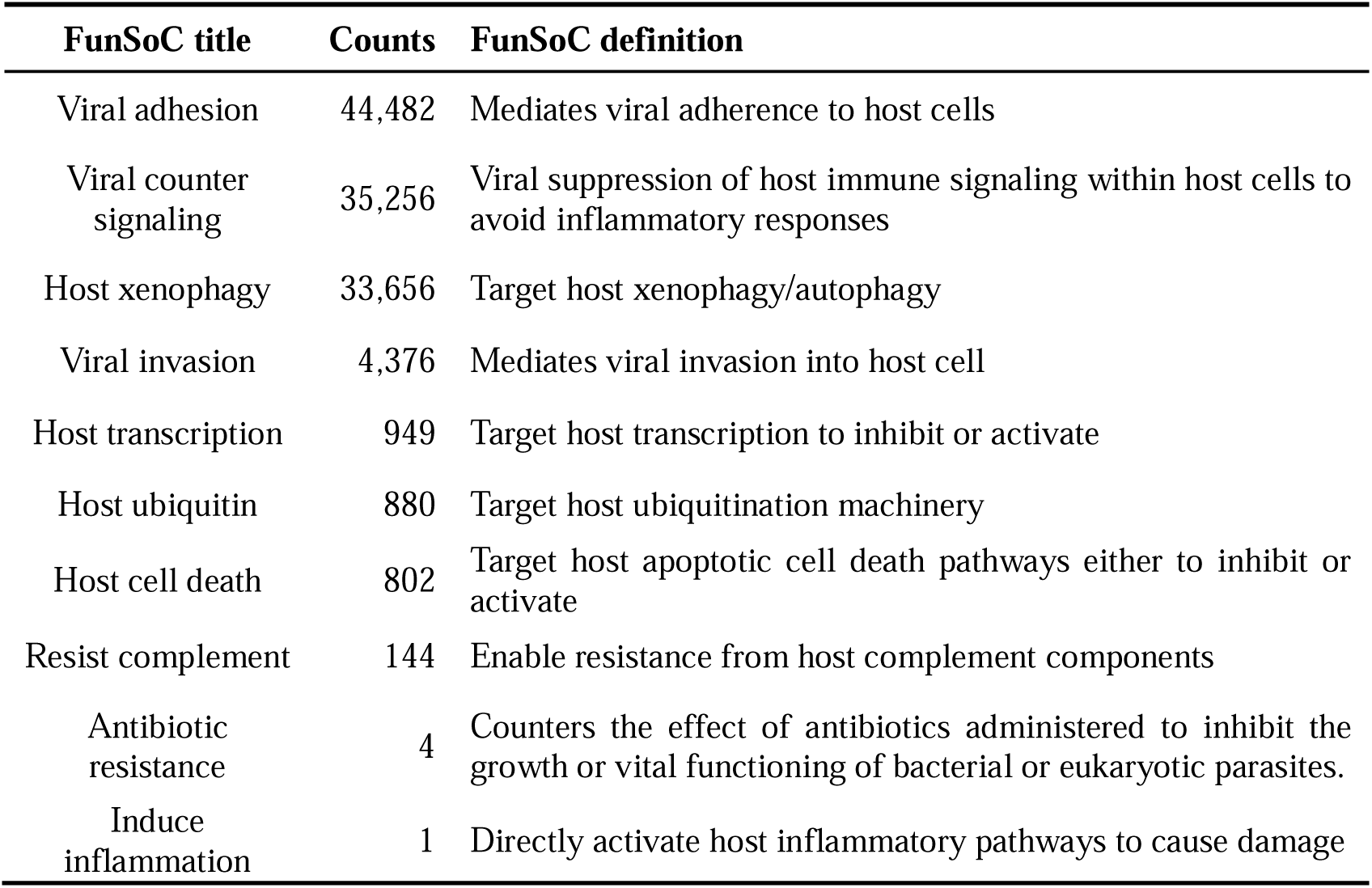
Pathogenicity functional information of viral sequences. Counts and definitions for ten identified functional signatures of concerns (FunSoCs) identified in this dataset using SeqScreen.

| FunSoC title | Counts | FunSoC definition |
| --- | --- | --- |
| Viral adhesion | 44,482 | Mediates viral adherence to host cells |
| Viral counter signaling | 35,256 | Viral suppression of host immune signaling within host cells to avoid inflammatory responses |
| Host xenophagy | 33,656 | Target host xenophagy/autophagy |
| Viral invasion | 4,376 | Mediates viral invasion into host cell |
| Host transcription | 949 | Target host transcription to inhibit or activate |
| Host ubiquitin | 880 | Target host ubiquitination machinery |
| Host cell death | 802 | Target host apoptotic cell death pathways either to inhibit or activate |
| Resist complement | 144 | Enable resistance from host complement components |
| Antibiotic resistance | 4 | Counters the effect of antibiotics administered to inhibit the growth or vital functioning of bacterial or eukaryotic parasites. |
| Induce inflammation | 1 | Directly activate host inflammatory pathways to cause damage |

Despite only 6.1% of the viral sequences in our dataset exhibiting “Viral invasion” (4,376), this feature is predominant among viruses without established zoonotic potential, surpassing the prevalence of the three previously mentioned pathogenic functions. A significant portion of these viruses, identified as ISVs, are mainly transmitted by arthropod vectors. However, a subset of these viruses has also been detected in *Artiodactyla*, *Perissodactyla*, and *Aves* (Figure 3A). In contrast, for viruses with known zoonotic potential, “viral adhesion,” “Viral counter-signaling,” and “Host xenophagy” emerge as the most prevalent pathogenic features. Contrasting with the viruses with non-established zoonotic potential, these viruses are predominantly associated with *Aves*, *Artiodactyla*, and *Rodentia* (Figure 3B).

**Figure 3:**
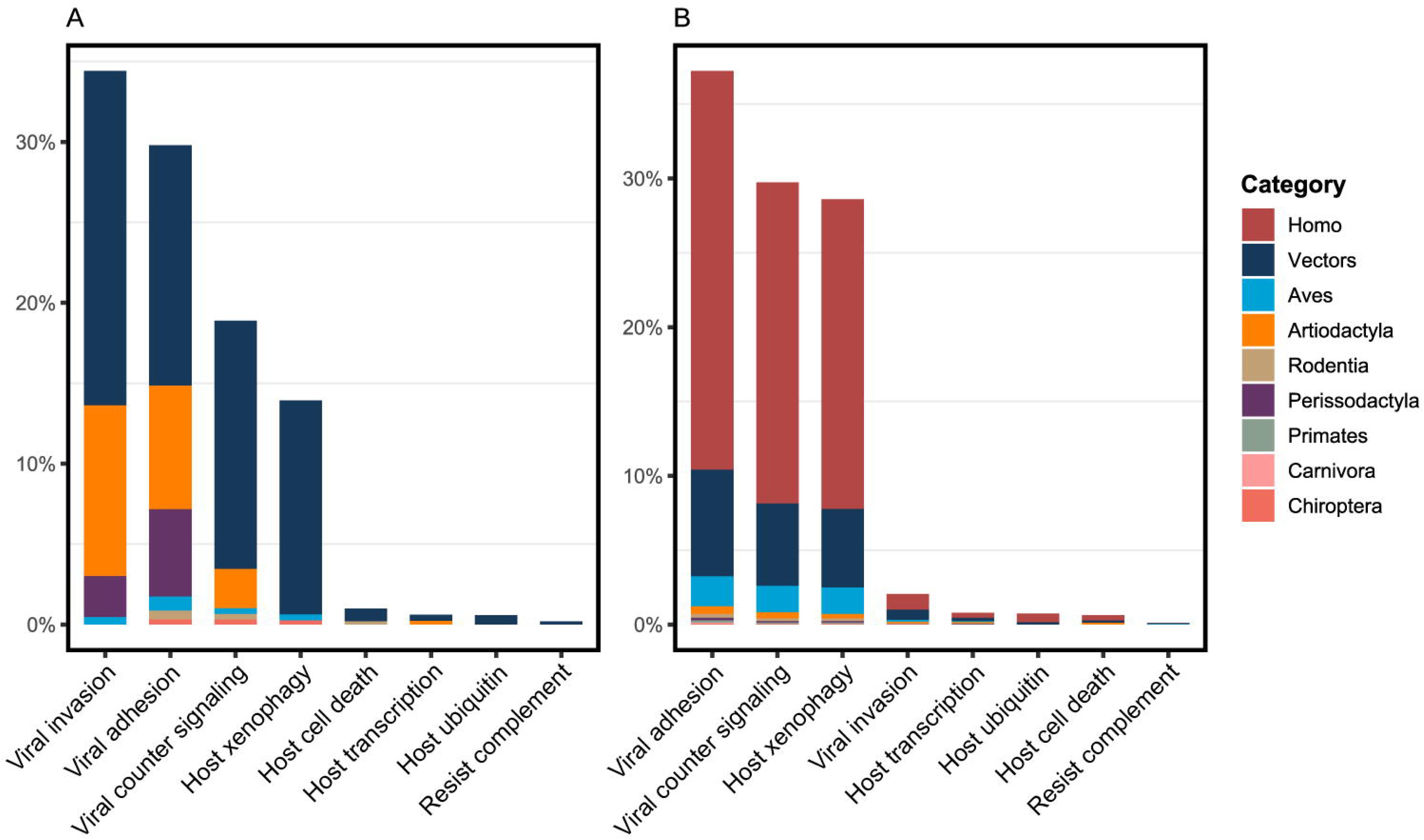
Distribution of host range across viruses with divergent pathogenic functions. The host range for viruses without zoonotic potential (A) and those possessing known zoonotic capabilities (B). The actual counts of viruses within each category are presented as percentage representations.

A binary classification model, employing the XGBoost algorithm with 11 viral features— including pathogenic features and the genomic size (length) of the virus—was developed. The model exhibited robust performance metrics: an accuracy of 94.87%, precision of 96.81%, recall of 96.77%, and an F1 score of 96.79%. Furthermore, the Receiver Operating Characteristic (ROC) curve and the confusion matrix provide comprehensive insights into the model’s performance (Figure 4).

**Figure 4:**
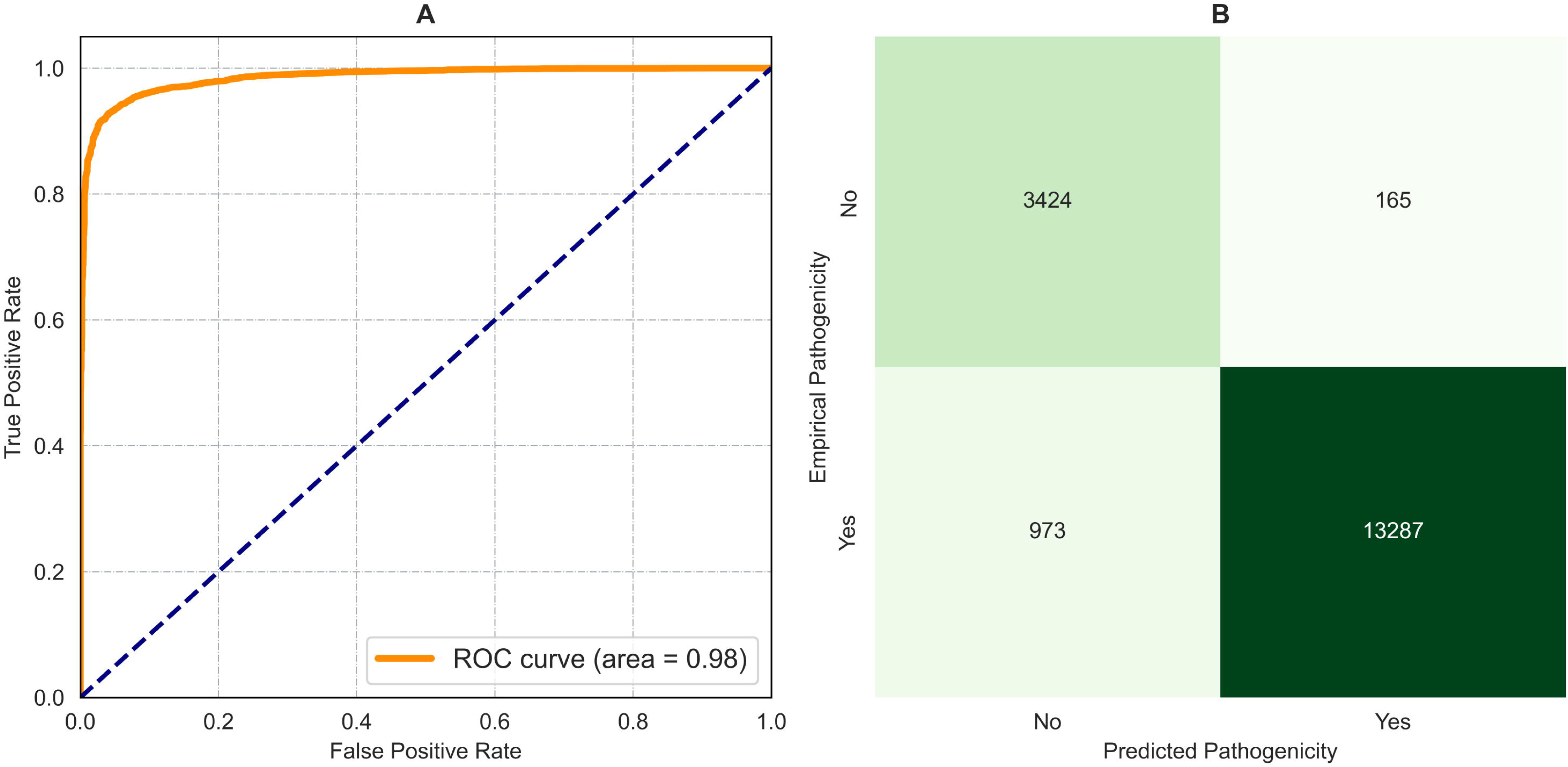
Comprehensive assessment of model performance metrics. Evaluating model performance through the utilization of the ROC Curve (A) and Confusion Matrix (B).

The evaluation of the classification model using test data reveals that “Viral adhesion” exhibits the highest average gain, highlighting its critical role in the model’s discernment of pathogenic potential. “Host xenophagy” and “Viral invasion” closely follow, each significantly influencing assessments of human pathogenicity (Figure 5A). In terms of average coverage, “Viral invasion” and “Host ubiquitin” stand out as the most influential features, signifying their impact on the virus pathogenicity (Figure 5B). Further examination of the model’s weights demonstrates the significance of the genomic size of the viral sequence, highlighting its vital contribution to refining pathogenicity assessments (Figure 5C). SHAP (SHapley Additive exPlanations) was utilized to perform an in-depth analysis of the contributions of individual pathogenic features to human pathogenicity of viruses. Notably, the feature with the highest ranking, “Viral size”, exhibited variable impacts on human pathogenicity, lacking a consistent pattern. In contrast, “Viral adhesion” and “Host xenophagy,” while slightly less influential than “Viral size,” demonstrated a strong tendency to increase the likelihood of a virus being pathogenic to humans. Conversely, “Viral invasion” was associated with a decreased likelihood of contributing to human pathogenicity, indicating an inverse effect. Furthermore, other pathogenic features, such as “Host transcription” and “Host ubiquitin” were found to positively correlate with pathogenicity (Figure 6).

**Figure 5:**
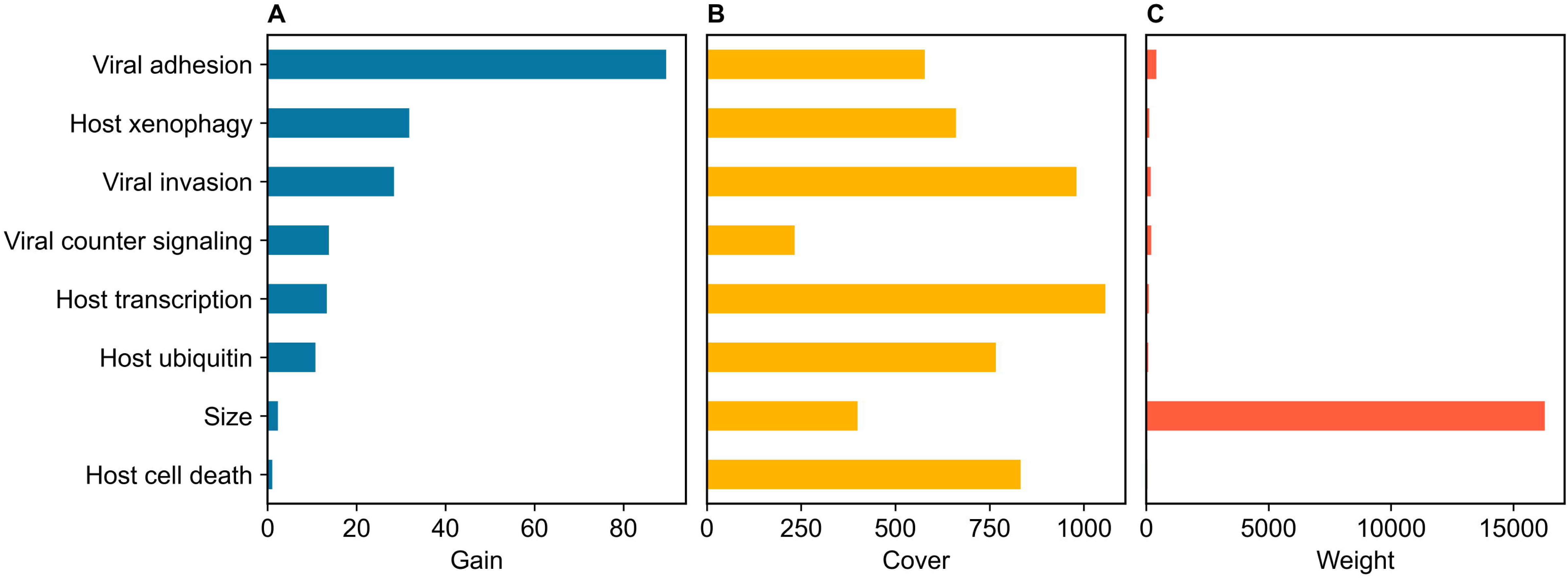
Comparative analysis of feature importance in pathogenicity assessment. In the evaluation of the XGBoost model, the importance of pathogenicity features is determined through three distinct metrics: Gain (A), Cover (B), and Weight (C). These metrics collectively illuminate the relative importance of each feature in assessing the human pathogenicity of viruses transmitted by hematophagous arthropod vectors.

**Figure 6:**
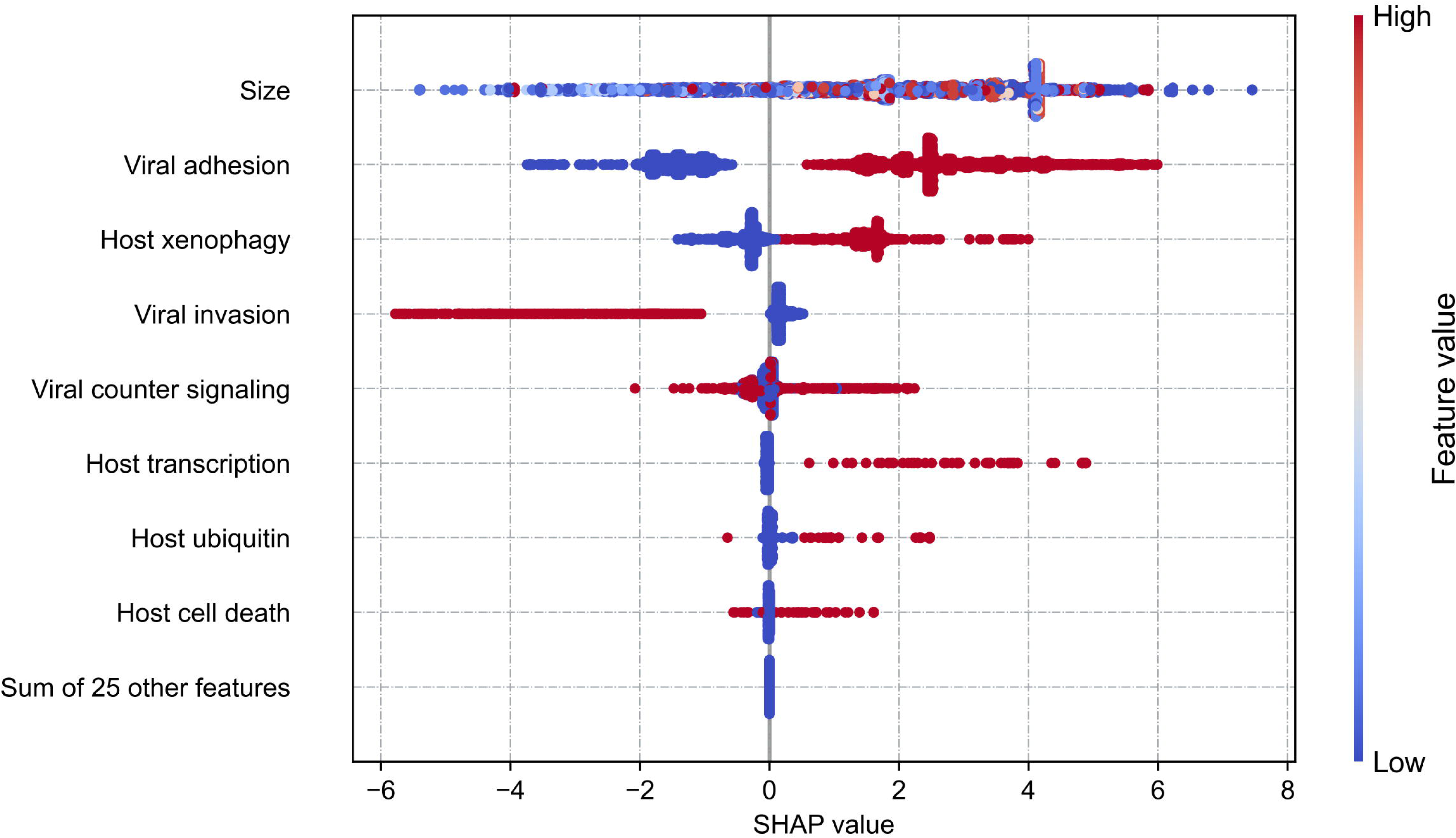
Impact of viral function on pathogenicity predictions via SHAP analysis. The collective contribution of various viral functions on the model’s pathogenicity predictions, as analyzed through SHAP.

Our interactive analysis delves into the complex interplay among pathogenic features. Specifically, a pronounced interaction between “Viral size” and “Viral counter signaling” was observed. However, this interaction does not present a clear trend regarding its impact on pathogenicity (Figure 7A). Furthermore, the coexistence of “Host xenophagy” and “Viral adhesion” within viral sequences significantly augments the likelihood of pathogenicity to humans, highlighting the critical role of their synergy in enhancing pathogenic potential (Figure 7B). Contrarily, “Viral invasion” and “Viral counter signaling” demonstrate a distinct interaction. While the presence of “Viral counter signaling” alone may suggest potential pathogenicity, the simultaneous presence of these two functions markedly decreases its human pathogenicity (Figure 7C). Similarly, while the present of “host xenophagy” along may increase the likelihood of viral pathogenicity, its concurrent occurrence with “viral adhesion” or “viral counter signaling” significantly mitigates this pathogenic potential (Figure s3).

**Figure 7:**
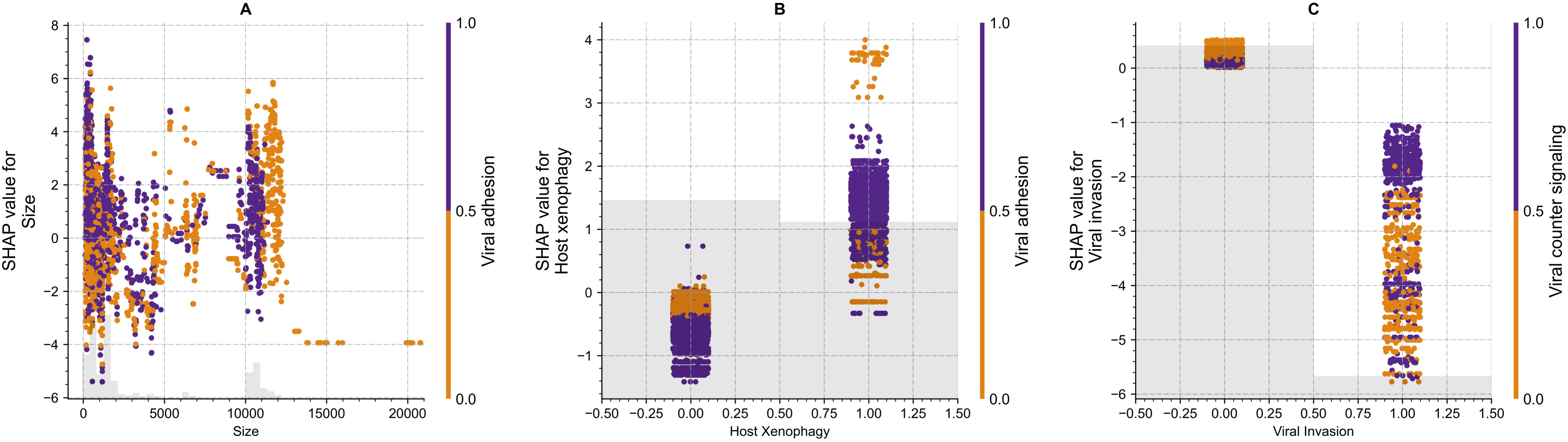
Interactions among key features in pathogenicity predictions. The specific interactions among critical features within pathogenicity prediction models, including the relationship between viral sequence size and Viral adhesion (A), Host xenophagy and Viral adhesion (B), as well as Viral invasion and Viral counter signaling (C).

### Predicting pathogenicity in external datasets using machine learning

This study utilized an external validation dataset of 228 viral sequences to evaluate the zoonotic potential of viruses using two machine learning models. The first model analyses host range features encoded in virus genomes to identify those with zoonotic potential. The second model, Arbovirus Human Pathogen Prediction (AHPP), aims to detect viruses that are pathogenic to humans by focusing on sequence functional information.

Predictive analysis of the zoonotic risk associated with all 64 distinct types of arboviruses and ISVs in the dataset revealed that 11 viruses were classified as having a “Very High” risk of zoonosis, 32 were “High”, and the remaining 21 were categorized as “Medium” and “Low”, indicating a lower likelihood of emerging as zoonotic pathogens (Supplementary Table 4). The AHPP model identified 11 strains from 5 viruses as potentially pathogenic to humans. In our database, among the viruses already known to affect humans, one strain of JEV and Bunyamwera virus (BUNV), 8 of 25 strains of Severe fever with thrombocytopenia syndrome virus (SFTSV) were predicted to be pathogenic to humans. Our model indicates that, despite being classified potentially as possible arboviruses, Palma virus and Zaliv Terpeniya virus (ZTV) may have the capacity to infect humans. The lack of current research confirming the ability of these viruses to infect humans highlights a possible gap in our understanding of their zoonotic risk. There are significant differences in the evaluations of pathogenicity for Ebinur Lake virus (EBIV) between the two models. Classified within the *Orthobunyavirus* genus, EBIV was assigned a “Very High” zoonotic potential rank by the first model. In contrast, the AHPP analysis of all 25 EBIV sequences in this dataset did not identify any pathogenic features, suggesting a low likelihood of EBIV posing a pathogenic threat to humans. Additionally, the AHPP model’s analysis revealed that the five strains of Nairobi sheep disease virus (NSDV), three strains of Restan virus (RESV), and Tataguine virus (TATV) present in this dataset exhibit no pathogenic features, thereby suggesting a diminished probability of these viruses infecting humans and causing disease. However, previous detections of NSDV, RESV, and TATV in human blood samples have suggested a considerable zoonotic potential for these viruses. The Zoonotic Rank model also highlight both RESV and TATV as having high zoonotic potential, indicating a discernible discrepancy between empirical observations and the model’s predictions.

## Discussion

The intricate interactions between hematophagous arthropods and the pathogenic viruses they harbor form a dynamic ecosystem with profound implications for public health. In recent decades, diseases transmitted by mosquitoes and ticks, such as ZIKAV and Jingmen tick virus (JMTV), have had significant social and economic impacts, underscoring the importance of monitoring and preventing the spread of potentially pathogenic viruses (Y.-J. S. Huang, Higgs, and Vanlandingham 2019b; Wu et al. 2023). Consequently, we have collected global data on vector-borne viruses from hematophagous arthropods and have developed XGBoost regression and classification prediction models to assess the pathogenicity of arboviruses and ISVs. This integration overcomes challenges associated with the traditional virological methods of virus isolation and cultivation. It facilitates the identification of key characteristics influencing viral pathogenicity and enhances our ability to predict viruses with potential zoonotic risks from viral sequences, thereby optimizing public health strategies and preventive measures.

In our dataset, the majority of arboviruses and ISVs are RNA viruses, predominantly consisting of dsRNA viruses categorized under Baltimore Group III. The distribution of these viruses is influenced by various factors; a detailed correlation analysis revealed significant correlations between most viruses and six key characteristics. These characteristics include the vector family and weather conditions, among others. Furthermore, within the community of vector-borne viruses, families such as *Flaviviridae*, *Togaviridae*, *Bunyaviridae*, *Rhabdoviridae*, and *Phenuiviridae* frequently co-occur and are considered part of the core virome (Coatsworth et al. 2022) (Figure supplement 2). Within the dynamic cycles of arboviruses and ISVs, numerous factors influence their transmission to humans and pathogenicity. Viral interactions with non-vector hosts are important drivers of viral evolution and are primarily responsible for zoonotic spillovers (Y.-J. S. Huang, Higgs, and Vanlandingham 2019a; Sen, Nayak, and De 2016). Research indicates that vectors’ interaction with avian hosts facilitates long-distance transmission, while interactions with vertebrates are pivotal determinants of spillovers (Forrester, Coffey, and Weaver 2014; García-Romero et al. 2023; Stephenson et al. 2019). Our findings reveal that non-vector hosts significantly affect human pathogenicity, especially within *Perissodactyla* and *Carnivora*, where a high abundance of vector-borne viruses correlates with increased human pathogenicity. The characteristics of vector hosts are also essential in this context. A higher diversity of viruses within vectors may lead co-infections, thereby facilitating viral evolution and spillovers (Vogels et al. 2019). Hematophagous vectors and environmental changes further impact viral transmission and pathogenicity (Hermanns et al. 2023; Weissenböck et al. 2010). Moreover, the inherent characteristics of viruses influence their ability to cause disease. Specifically, most members of the *Flaviviridae* family are recognized as pathogenic to humans (Zhenzhen Zhang, Rong, and Li 2019).

Given that the genetic functionalities of viruses can predict their pathogenicity (Bartoszewicz, Seidel, and Renard 2021), in this study, we developed another XGBoost algorithm-based classification model, termed AHHP, to predict human pathogenicity based on the genetic functionalities of arboviruses and ISVs. The AHHP demonstrates superior performance, highlighting the significant contributions of individual viral functions to pathogenicity. Specifically, “Viral adhesion”, a crucial process facilitating viral infection and entry into host cells, markedly increases the pathogenicity of viruses to humans. For example, DENV, WNV and ZIKAV from *Flaviviridae* family utilize their envelope (E) and capsid proteins to engage with receptor cells (Begum et al. 2019; Faustino et al. 2019; Cruz-Oliveira et al. 2015; Martins et al. 2019; Hasan et al. 2017). Similarly, the Chikungunya virus (CHIKV) from the *Togaviridae* family promotes fusion with receptor cells through its trimeric E1/E2 spikes (Ciota and Keyel 2019). Although “Viral invasion” serves a crucial role similar to “Viral adhesion” in the initial phase of viral entry, this feature is less prevalent in our dataset, appearing primarily in arboviruses and ISVs that are not currently pathogenic to humans. Despite efforts to balance the positive and negative samples during the training process, it was challenging to completely eliminate this influence. As a result, this feature was identified as a protective factor against viral pathogenicity in humans in our study. The interaction of this feature with “Viral counter signaling” also suggests that its presence significantly inclines the virus towards non-pathogenicity. However, in the actual process of viral infection, these two processes can coexist. Therefore, these findings may indicate that the pathogenic mechanisms of vector-borne viruses could involve unique mechanisms. The function “Host xenophagy” and “Host transcription” enable viruses to maintain a prolonged presence within host cells, helping to facilitate the exploitation of cellular resources for replication and spread (King, Wegman, and Endy 2020). The feature “size” is not directly associated with pathogenic functions but is crucial for refining prediction outputs. Training the model using only 33 functional features yielded an unreliable accuracy of 82% and a high false-positive rate. Including “size” significantly enhanced model performance. However, the influence of “size” on viral pathogenicity exhibits a random distribution trend, and interaction analysis also shows that its effect with “Viral counter signaling” lacks clear directionality. Overall, “size” fine-tunes the model’s predictions, and its integration with functional features leads to a more accurate assessment of the likelihood of pathogenicity to humans.

Hematophagous arthropod vector-borne viruses collected after 2022 were used as an external validation dataset to assess the predictive accuracy of the AHHP model, comparing it with another model, the Zoonotic Rank. Both models accurately identified the known arboviruses—SFTS, JEV, and BUNV—as strongly pathogenic to humans. All 25 strains of the SFTS virus were sourced from ticks in Miyazaki Prefecture, Japan, a region previously affected by two confirmed SFTS cases (Sato et al. 2021). Phylogenetic analysis revealed a high homology with a virus isolated from an SFTS patient, confirming a close genetic relationship. Similarly, a strain of JEV was isolated from mosquitoes in the Qinghai-Tibet Plateau of China (Li et al. 2011). Despite the region’s high altitude, the detection of antibodies in both the indigenous population and swine indicates localized virus transmission. BUNV was sourced from *Aedes ochraceus* mosquitoes in Desai, Garissa, Kenya (Kapuscinski et al. 2021), and variants of this virus have been associated with large-scale outbreaks of african hemorrhagic fever (Gerrard et al. 2004). Given that BUNV can be transmitted through water, particular attention is warranted to its potential spread by female mosquitoes during oviposition (Turner and Christofferson 2024). In addition to well-recognized pathogenic, the AHHP model predicted ZTV and Palma virus as potentially highly pathogenic arboviruses. ZTV was isolated from *Ixodes putus* and has been found in *Culex modestus*, suggesting potential cross-vector host transmission (Kapuscinski et al. 2021). ZTV and SFTSV both belong to the *Bandavirus* genus. While the public health significance of ZTV is currently unclear, our research supports its classification as an arbovirus, and further pathogenicity testing is recommended (Palacios et al. 2013). Conversely, despite the Zoonotic Rank model considering it not to pose zoonotic potential, the AHHP model identified the Palma virus as potentially highly pathogenic based on its “Viral adhesion” and “Host cell death” features. However, there are some discrepancies between the predictions of the AHHP model and the Zoonotic Rank model. For example, EBIV, isolated from *Culex modestus*, can infect BALB/c mice and cause pronounced clinical symptoms (Zhao et al. 2020); it has also been shown that *Aedes aegypti* and *Hydrocoloeus minutus* can carry this virus, with antibodies detected in human serum samples (C. Yang et al. 2022). Yet, the absence of positive RT-PCR results currently hampers confirmation of its ability to infect and induce disease in human (Xia et al. 2020). Furthermore, none of the 25 EBIV sequences analyzed in this study exhibited pathogenic functional, leading to their classification as non-pathogenic to humans. This suggests that further empirical research is necessary to verify the pathogenicity of arboviruses. Notably, the predictive results of the AHHP model diverged from known clinical observations in some instances. For example, the Nairobi Sheep Disease virus (NSDV) and Restan virus (RESV), both previously detected in humans, were classified as non-pathogenic in this study because no pathogenic functional were detected.

In addition, our study has certain limitations. The dataset of global viruses used exhibits an uneven distribution, with certain viruses being overrepresented. This imbalance could introduce an unavoidable bias, potentially affecting the accuracy of our model. Additionally, our pathogenicity training, which is focused solely on the pathogenic functional features of viral sequences, may overlook other important features related to their pathogenicity. Also, variations in blood-feeding preferences among different hematophagous vectors can significantly influence both the transmission and pathogenicity of viruses. To enhance the robustness of future studies, it is crucial to incorporate a broader range of data that includes these behavioral nuances.

In summary, our study identified the principal epidemiological factors influencing the pathogenicity of hematophagous arthropod vector-borne viruses. A machine learning predictive model was developed, revealing key features associated with viral pathogenicity and enabling the prediction of pathogenicity at the viral strain level. This model was evaluated alongside the Zoonotic Rank model using an additional validation dataset, demonstrating its reliability. We aim to apply this model more broadly to metagenomic results to explore potential pathogenic viruses within extensive data on hematophagous arthropod vectors. Such application could help mitigate current and future risks associated with vector-borne diseases.

**Table 2:** Zoonotic risk assessment for arboviruses using two models in an external validation dataset. The classification, accession numbers, arbovirus status, pathogenic functional features, and zoonotic status of viruses. “Risk Rank” is determined using the Zoonotic Risk Assessment tool, while “AHPP” refers to the Arbovirus Human Pathogen Prediction results developed in this study. VCS: Viral counter signaling; HX: Host xenophagy; HCD: Host cell death; VA: Viral adhesion.

| Family | Genus | Virus | Accession | Status | VA | HX | VSC | HCD | Known Zoonotic | Risk Rank | AHPP |
| --- | --- | --- | --- | --- | --- | --- | --- | --- | --- | --- | --- |
| Flaviviridae | Orthoflavivirus | Japanese encephalitis virus (JEV) | HQ652538 | Arbovirus | 1 | 1 | 1 | 0 | 1 | Very high | 1 |
| Nairoviridae | Orthonairovirus | Nairobi sheep disease virus (NSDV) | MZ244233 | Arbovirus | 0 | 0 | 0 | 0 | 1 | Medium | 0 |
| Peribunyaviridae | Orthobunyavirus | Bunyamwera virus (BUNV) | MH484289 | Arbovirus | 1 | 0 | 0 | 0 | 1 | High | 1 |
|  |  | Ebinur Lake virus (EBIV) | NC_079003-NC_079005<br>OR861624<br>OM800984<br>OM800987-OM800991<br>OM800994<br>OM800996-OM800998<br>OM801001-OM801005<br>OM801007 | - | 0 | 0 | 0 | 0 | 0 | Very high | 0 |
|  |  | Restan virus (RESV) | MK896475-MK896477 | Arbovirus | 0 | 0 | 0 | 0 | 1 | Very high | 0 |
|  |  | Tataguine virus (TATV) | MK896454-MK896456 | Probable Arbovirus | 0 | 0 | 0 | 0 | 1 | High | 0 |
| Phenuiviridae | Bandavirus | Palma virus | MK896493 | - | 1 | 0 | 0 | 1 | 0 | Medium | 1 |
|  |  | Severe fever with thrombocytopenia syndrome virus (SFTSV) | LC536536 -<br>LC536542<br>LC536546 -<br>LC536552<br>LC536556 -<br>LC536562 | Arbovirus | 1 | 0 | 0 | 1 | 1 | Very high | 1 |
|  | Uukuvirus | Zaliv Terpeniya virus (ZTV) | MK896425 | Possible Arbovirus | 1 | 0 | 0 | 0 | 0 | High | 1 |

## Data Availability

The raw dataset supporting the findings of this study is available on Figshare at https://doi.org/10.6084/m9.figshare.22154573.v5. Additionally, the processed data used and/or analyzed during the current study can be obtained by contacting the corresponding author. Requests for access to the processed data will be promptly addressed.

## Acknowledgement

The authors acknowledge the global open dataset shared by Huang et al and Xuan Li for assistance with additional data collection. The laboratory is funded by a grant from National Key Research and Development Program of China (2019YFC1200501).

## Data availability

The data supporting the findings of this study are available upon reasonable request from the author. Researchers interested in accessing the dataset for further exploration or verification are encouraged to contact Huakai Hu at for assistance. We are committed to promoting transparency and collaboration in scientific research, and we welcome inquiries regarding the data underlying our published results.

## Author contribution

Huakai Hu, Idea Generation, Data Curation and Transformation, Model Development and validation, Visualization, Writing – original draft, review and editing; Chaoying Zhao, Conceptualization, Methodology, Writing – review and editing; Jiali Chen, Conceptualization, Methodology, Writing – review and editing; Meiling Jin, Conceptualization, Writing – review and editing; Hua Shi, Conceptualization, Writing – review and editing; Jinpeng Guo, Project administration, Writing – review and editing; Changjun Wang, Conceptualization, Methodology, Writing – review and editing; Yong Chen, Supervision, Funding acquisition, Project administration, Writing – review and editing;

## Supplementary Information

**Supplementary Table 1:**
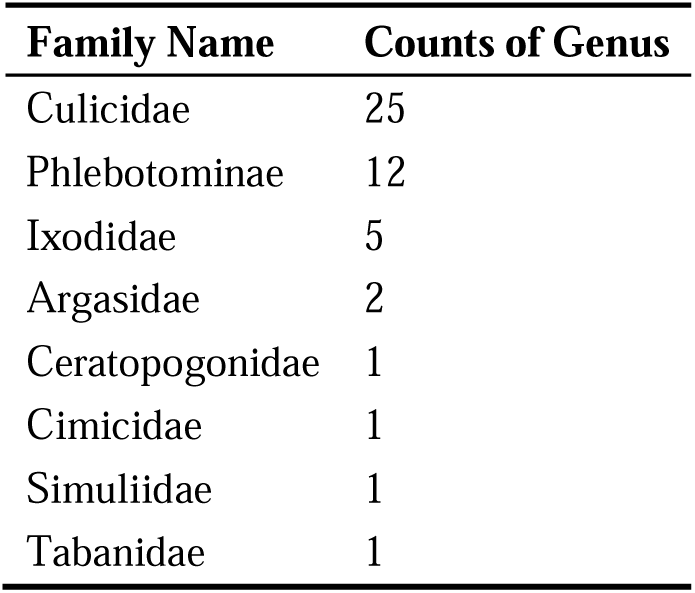
Family and genus of hematophagous arthropod vector in database. The family of of hematophagous arthropods examined and enumerates the genera within each family.

**Supplementary Table 2:**
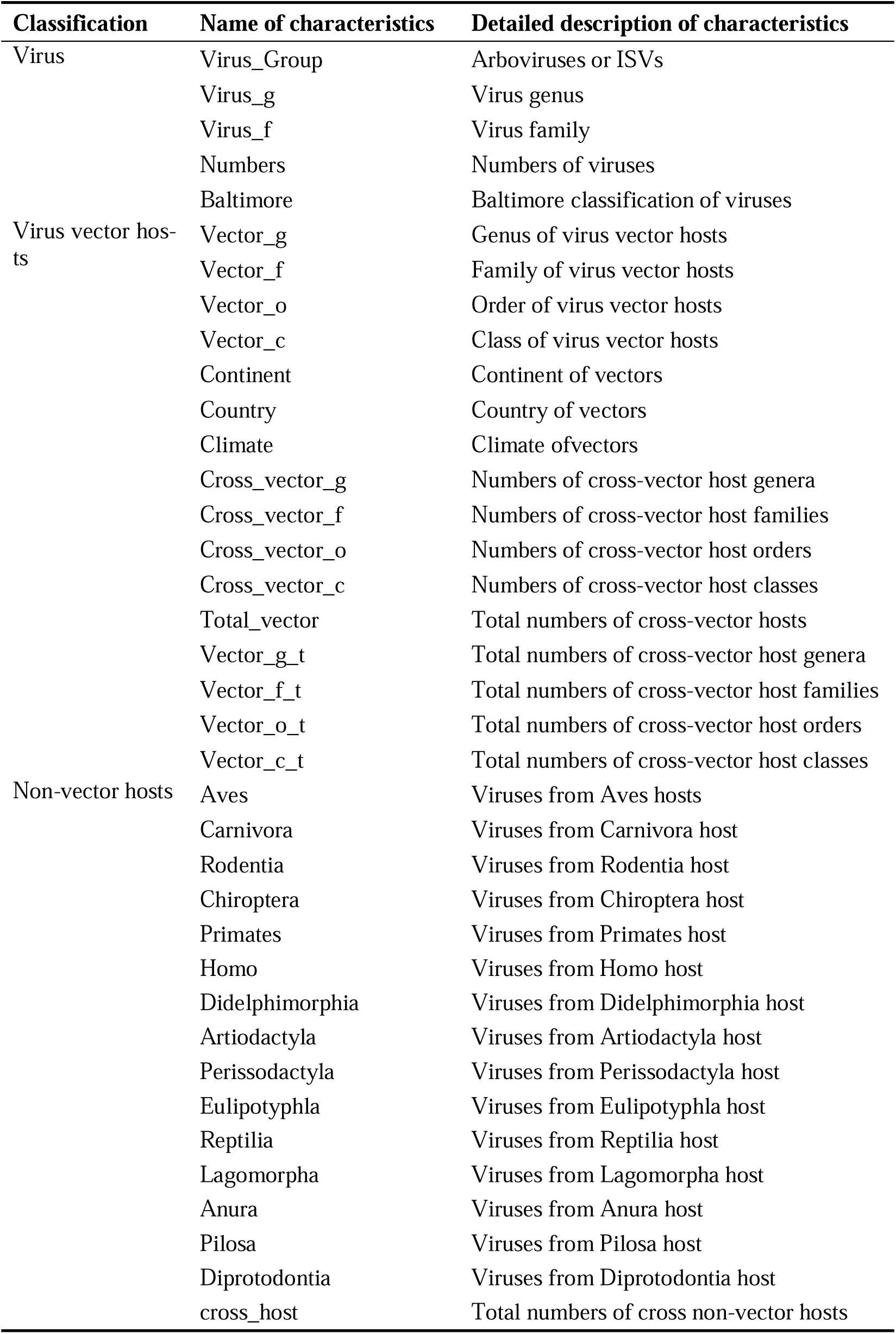
Epidemiological characteristics in regression model. A summary of the 37 variables, categorized into three groups, utilized in our regression model.

| Classification | Name of characteristics | Detailed description of characteristics |
| --- | --- | --- |
| Virus | Virus_Group | Arboviruses or ISVs |
|  | Virus_g | Virus genus |
|  | Virus_f | Virus family |
|  | Numbers | Numbers of viruses |
|  | Baltimore | Baltimore classification of viruses |
| Virus vector hosts | Vector_g | Genus of virus vector hosts |
|  | Vector_f | Family of virus vector hosts |
|  | Vector_o | Order of virus vector hosts |
|  | Vector_c | Class of virus vector hosts |
|  | Continent | Continent of vectors |
|  | Country | Country of vectors |
|  | Climate | Climate of vectors |
|  | Cross_vector_g | Numbers of cross-vector host genera |
|  | Cross_vector_f | Numbers of cross-vector host families |
|  | Cross_vector_o | Numbers of cross-vector host orders |
|  | Cross_vector_c | Numbers of cross-vector host classes |
|  | Total_vector | Total numbers of cross-vector hosts |
|  | Vector_g_t | Total numbers of cross-vector host genera |
|  | Vector_f_t | Total numbers of cross-vector host families |
|  | Vector_o_t | Total numbers of cross-vector host orders |
|  | Vector_c_t | Total numbers of cross-vector host classes |
| Non-vector hosts | Aves | Viruses from Aves hosts |
|  | Carnivora | Viruses from Carnivora host |
|  | Rodentia | Viruses from Rodentia host |
|  | Chiroptera | Viruses from Chiroptera host |
|  | Primates | Viruses from Primates host |
|  | Homo | Viruses from Homo host |
|  | Didelphimorphia | Viruses from Didelphimorphia host |
|  | Artiodactyla | Viruses from Artiodactyla host |
|  | Perissodactyla | Viruses from Perissodactyla host |
|  | Eulipotyphla | Viruses from Eulipotyphla host |
|  | Reptilia | Viruses from Reptilia host |
|  | Lagomorpha | Viruses from Lagomorpha host |
|  | Anura | Viruses from Anura host |
|  | Pilosa | Viruses from Pilosa host |
|  | Diprotodontia | Viruses from Diprotodontia host |
|  | cross_host | Total numbers of cross non-vector hosts |

**Supplementary Table 3:** Hyperparameter settings for the XGBoost regression model. Optimized parameter settings for the XGBoost regression model obtained through experimentation.

| Hyperparameter | Value |
| --- | --- |
| booster | dart |
| eta | 0.15 |
| max_depth | 3 |
| subsample | 0.7 |
| objective | reg:logistic |
| tree_method | exact |
| max_cat_threshold | 20 |
| scale_pos_weight | 0.792 |
| eval_metric | ["logloss", "auc", 'error'] |

**Supplementary Table 4:** Hyperparameter settings for the XGBoost classification model. Optimized parameter settings for the XGBoost classification model obtained through experimentation.

| Hyperparameter | Value |
| --- | --- |
| objective | binary:logistic |
| tree_method | exact |
| scale_pos_weight | 0.26 |
| eta | 0.15 |

**Supplementary Table 5:** Assessment Results of Zoonotic Potential for Arboviruses in the External Validation Dataset. The 95% confidence intervals for scores delineate the range of values for assessing zoonotic potential. In this zoonotic ranking system, the classifications of “Very High” and “High” indicate a significant zoonotic potential, while “Medium” and “Low” classifications suggest a reduced zoonotic potential, indicating a lower probability of human infection.

| **Virus** | **Score (95%CI)** | **Zoonotic Rank** |
| --- | --- | --- |
| Aino virus | 0.7 [0.37, 0.93] | Very high |
| Antequera virus | 0.54 [0.32, 0.84] | Very high |
| Birao virus | 0.65 [0.38, 0.92] | Very high |
| Ebinur lake virus | 0.53 [0.33, 0.78] | Very high |
| Japanese encephalitis virus | 0.62 [0.37, 0.87] | Very high |
| Kaikalur virus | 0.61 [0.32, 0.85] | Very high |
| Potosi virus | 0.61 [0.38, 0.86] | Very high |
| Restan virus | 0.56 [0.35, 0.82] | Very high |
| Severe fever with thrombocytopenia syndrome virus | 0.62 [0.32, 0.9] | Very high |
| Termeil virus | 0.56 [0.33, 0.85] | Very high |
| Turlock orthobunyavirus | 0.48 [0.3, 0.81] | Very high |
| Abras virus | 0.49 [0.29, 0.71] | High |
| Barranqueras virus | 0.38 [0.22, 0.54] | High |
| Bertioga virus | 0.35 [0.18, 0.55] | High |
| Boraceia virus | 0.35 [0.2, 0.58] | High |
| Bozo virus | 0.39 [0.22, 0.6] | High |
| Bruconha virus | 0.32 [0.15, 0.54] | High |
| Bunyamwera virus | 0.46 [0.24, 0.73] | High |
| Caimito virus | 0.46 [0.26, 0.75] | High |
| Enseada virus | 0.33 [0.17, 0.55] | High |
| Guaratuba virus | 0.33 [0.19, 0.58] | High |
| Gumbo Limbo virus | 0.43 [0.24, 0.67] | High |
| Hughes orthonairovirus | 0.31 [0.17, 0.55] | High |
| Kaisodi virus | 0.39 [0.21, 0.59] | High |
| Las Maloyas virus | 0.39 [0.21, 0.59] | High |
| Nola virus | 0.33 [0.16, 0.53] | High |
| Northway virus | 0.32 [0.15, 0.54] | High |
| Okola virus | 0.41 [0.24, 0.61] | High |
| Oya virus | 0.3 [0.16, 0.55] | High |
| Peaton virus | 0.4 [0.24, 0.58] | High |
| Pueblo Viejo virus | 0.3 [0.15, 0.51] | High |
| Resistencia virus | 0.48 [0.27, 0.77] | High |
| Shokwe virus | 0.31 [0.16, 0.48] | High |
| Sunday Canyon virus | 0.35 [0.18, 0.58] | High |
| Tataguine virus | 0.36 [0.22, 0.66] | High |
| Tensaw virus | 0.46 [0.22, 0.75] | High |
| Tinaroo virus | 0.4 [0.23, 0.69] | High |
| Tlacotalpan virus | 0.52 [0.28, 0.8] | High |
| Wanowrie virus | 0.31 [0.16, 0.55] | High |
| Wongal virus | 0.3 [0.16, 0.51] | High |
| Wyeomyia_orthobunyavirus | 0.46 [0.25, 0.71] | High |
| Yaba-7 virus | 0.34 [0.17, 0.58] | High |
| Zaliv Terpeniya virus | 0.51 [0.28, 0.78] | High |
| Anopheles B virus | 0.24 [0.13, 0.42] | Medium |
| Bushbush virus | 0.27 [0.12, 0.44] | Medium |
| Cacao virus | 0.19 [0.09, 0.35] | Medium |
| Estero Real orthobunyavirus | 0.17 [0.09, 0.32] | Medium |
| Gamboa virus | 0.26 [0.15, 0.4] | Medium |
| Huangpi Tick Virus 1 | 0.21 [0.1, 0.34] | Medium |
| Kairi virus | 0.29 [0.13, 0.55] | Medium |
| Ketapang virus | 0.28 [0.14, 0.47] | Medium |
| Lednice virus | 0.24 [0.13, 0.43] | Medium |
| MAG: Nairobi sheep disease virus | 0.22 [0.09, 0.39] | Medium |
| Main Drain virus | 0.18 [0.08, 0.34] | Medium |
| Moriche virus | 0.28 [0.13, 0.45] | Medium |
| Pahayokee virus | 0.22 [0.12, 0.34] | Medium |
| Palma virus | 0.29 [0.15, 0.5] | Medium |
| San Juan virus | 0.25 [0.14, 0.39] | Medium |
| Santa Rosa virus | 0.24 [0.12, 0.44] | Medium |
| Sapphire II virus | 0.2 [0.1, 0.32] | Medium |
| Tanga virus | 0.24 [0.14, 0.41] | Medium |
| Balsa almendravirus | 0.15 [0.08, 0.24] | Low |
| Coot Bay virus | 0.15 [0.07, 0.24] | Low |
| Estero Real virus | 0.18 [0.1, 0.29] | Low |

## References

Balaji A, Kille B, Kappell AD, Godbold GD, Diep M, Elworth RAL, Qian Z, Albin D, Nasko DJ, Shah N, Pop M, Segarra S, Ternus KL, Treangen TJ. 2022. SeqScreen: accurate and sensitive functional screening of pathogenic sequences via ensemble learning. Genome Biol 23:133. doi:10.1186/s13059-022-02695-x

Bartoszewicz JM, Genske U, Renard BY. 2021a. Deep learning-based real-time detection of novel pathogens during sequencing. Briefings in Bioinformatics 22:bbab269. doi:10.1093/bib/bbab269

Bartoszewicz JM, Seidel A, Renard BY. 2021b. Interpretable detection of novel human viruses from genome sequencing data. NAR Genomics and Bioinformatics 3:lqab004. doi:10.1093/nargab/lqab004

Batson J, Dudas G, Haas-Stapleton E, Kistler AL, Li LM, Logan P, Ratnasiri K, Retallack H. 2021. Single mosquito metatranscriptomics identifies vectors, emerging pathogens and reservoirs in one assay. eLife 10:e68353. doi:10.7554/eLife.68353

Begum F, Das S, Mukherjee D, Ray U. 2019. Hijacking the Host Immune Cells by Dengue Virus: Molecular Interplay of Receptors and Dengue Virus Envelope. Microorganisms 7:323. doi:10.3390/microorganisms7090323

Behl A, Nair A, Mohagaonkar S, Yadav P, Gambhir K, Tyagi N, Sharma RK, Butola BS, Sharma N. 2022. Threat, challenges, and preparedness for future pandemics: A descriptive review of phylogenetic analysis based predictions. Infect Genet Evol 98:105217. doi:10.1016/j.meegid.2022.105217

Birnberg L, Temmam S, Aranda C, Correa-Fiz F, Talavera S, Bigot T, Eloit M, Busquets N. 2020. Viromics on Honey-Baited FTA Cards as a New Tool for the Detection of Circulating Viruses in Mosquitoes. Viruses 12:274. doi:10.3390/v12030274

Brinkmann A, Nitsche A, Kohl C. 2016. Viral Metagenomics on Blood-Feeding Arthropods as a Tool for Human Disease Surveillance. International Journal of Molecular Sciences 17:1743. doi:10.3390/ijms17101743

Calisher CH, Higgs S. 2018. The Discovery of Arthropod-Specific Viruses in Hematophagous Arthropods: An Open Door to Understanding the Mechanisms of Arbovirus and Arthropod Evolution? Annual Review of Entomology 63:87–103. doi:10.1146/annurev-ento-020117-043033

Ciota AT, Keyel AC. 2019. The Role of Temperature in Transmission of Zoonotic Arboviruses. Viruses 11:1013. doi:10.3390/v11111013

Coatsworth H, Bozic J, Carrillo J, Buckner EA, Rivers AR, Dinglasan RR, Mathias DK. 2022. Intrinsic variation in the vertically transmitted core virome of the mosquito Aedes aegypti. Molecular Ecology 31:2545–2561. doi:10.1111/mec.16412

Conway MJ, Colpitts TM, Fikrig E. 2014. Role of the Vector in Arbovirus Transmission. Annu Rev Virol 1:71–88. doi:10.1146/annurev-virology-031413-085513

Cruz-Oliveira C, Freire JM, Conceição TM, Higa LM, Castanho MARB, Da Poian AT. 2015. Receptors and routes of dengue virus entry into the host cells. FEMS Microbiology Reviews 39:155–170. doi:10.1093/femsre/fuu004

Cuthbert RN, Darriet F, Chabrerie O, Lenoir J, Courchamp F, Claeys C, Robert V, Jourdain F, Ulmer R, Diagne C, Ayala D, Simard F, Morand S, Renault D. 2023. Invasive hematophagous arthropods and associated diseases in a changing world. Parasites & Vectors 16:291. doi:10.1186/s13071-023-05887-x

Fang Z, Tan J, Wu S, Li M, Xu C, Xie Z, Zhu H. 2019. PPR-Meta: a tool for identifying phages and plasmids from metagenomic fragments using deep learning. GigaScience 8:giz066. doi:10.1093/gigascience/giz066

Faustino AF, Martins AS, Karguth N, Artilheiro V, Enguita FJ, Ricardo JC, Santos NC, Martins IC. 2019. Structural and Functional Properties of the Capsid Protein of Dengue and Related Flavivirus. International Journal of Molecular Sciences 20:3870. doi:10.3390/ijms20163870

Forrester NL, Coffey LL, Weaver SC. 2014. Arboviral Bottlenecks and Challenges to Maintaining Diversity and Fitness during Mosquito Transmission. Viruses 6:3991– 4004. doi:10.3390/v6103991

Fournet N, Voiry N, Rozenberg J, Bassi C, Cassonnet C, Karch A, Durand G, Grard G, Modenesi G, Lakoussan S-B, Tayliam N, Zatta M, Gallien S, investigation team,Noël H, Brichler S, Tarantola A. 2023. A cluster of autochthonous dengue transmission in the Paris region - detection, epidemiology and control measures, France, October 2023. Euro Surveill 28. doi:10.2807/1560-7917.ES.2023.28.49.2300641

García-Romero C, Carrillo Bilbao GA, Navarro J-C, Martin-Solano S, Saegerman C. 2023. Arboviruses in Mammals in the Neotropics: A Systematic Review to Strengthen Epidemiological Monitoring Strategies and Conservation Medicine. Viruses 15:417. doi:10.3390/v15020417

Geoghegan JL, Holmes EC. 2018. The phylogenomics of evolving virus virulence. Nat Rev Genet 19:756–769. doi:10.1038/s41576-018-0055-5

Gerrard SR, Li L, Barrett AD, Nichol ST. 2004. Ngari virus is a Bunyamwera virus reassortant that can be associated with large outbreaks of hemorrhagic fever in Africa. J Virol 78:8922–8926. doi:10.1128/JVI.78.16.8922-8926.2004

Gould E, Pettersson J, Higgs S, Charrel R, de Lamballerie X. 2017. Emerging arboviruses: Why today? One Health 4:1–13. doi:10.1016/j.onehlt.2017.06.001

Hasan SS, Miller A, Sapparapu G, Fernandez E, Klose T, Long F, Fokine A, Porta JC, Jiang W, Diamond MS, Crowe Jr. JE, Kuhn RJ, Rossmann MG. 2017. A human antibody against Zika virus crosslinks the E protein to prevent infection. Nat Commun 8:14722. doi:10.1038/ncomms14722

Hermanns K, Marklewitz M, Zirkel F, Kopp A, Kramer-Schadt S, Junglen S. 2023. Mosquito community composition shapes virus prevalence patterns along anthropogenic disturbance gradients. eLife 12:e66550. doi:10.7554/eLife.66550

Huang Y, Wang S, Liu H, Atoni E, Wang F, Chen W, Li Z, Rodriguez S, Yuan Z, Ming Z, Xia H. 2023. A global dataset of sequence, diversity and biosafety recommendation of arbovirus and arthropod-specific virus. Sci Data 10:305. doi:10.1038/s41597-023-02226-8

Huang Yan-Jang S., Higgs S, Vanlandingham DL. 2019. Arbovirus-Mosquito Vector-Host Interactions and the Impact on Transmission and Disease Pathogenesis of Arboviruses. Frontiers in Microbiology 10.

Huang Yan-Jang S, Higgs S, Vanlandingham DL. 2019. Emergence and re-emergence of mosquito-borne arboviruses. Current Opinion in Virology, Emerging viruses: intraspecies transmission • Viral Immunology 34:104–109. doi:10.1016/j.coviro.2019.01.001

Kampen H, Werner D. 2014. Out of the bush: the Asian bush mosquito Aedes japonicus japonicus (Theobald, 1901) (Diptera, Culicidae) becomes invasive. Parasit Vectors 7:59. doi:10.1186/1756-3305-7-59

Kapuscinski ML, Bergren NA, Russell BJ, Lee JS, Borland EM, Hartman DA, King DC, Hughes HR, Burkhalter KL, Kading RC, Stenglein MD. 2021. Genomic characterization of 99 viruses from the bunyavirus families Nairoviridae, Peribunyaviridae, and Phenuiviridae, including 35 previously unsequenced viruses. PLoS Pathog 17:e1009315. doi:10.1371/journal.ppat.1009315

Khongwichit S, Chuchaona W, Vongpunsawad S, Poovorawan Y. 2023. Molecular epidemiology, clinical analysis, and genetic characterization of Zika virus infections in Thailand (2020-2023). Sci Rep 13:21030. doi:10.1038/s41598-023-48508-4

King CA, Wegman AD, Endy TP. 2020. Mobilization and Activation of the Innate Immune Response to Dengue Virus. Frontiers in Cellular and Infection Microbiology 10.

Lewis WH, Tahon G, Geesink P, Sousa DZ, Ettema TJG. 2021. Innovations to culturing the uncultured microbial majority. Nat Rev Microbiol 19:225–240. doi:10.1038/s41579-020-00458-8

Li Y-X, Li M-H, Fu S-H, Chen W-X, Liu Q-Y, Zhang H-L, Da W, Hu S-L, La Mu SD, Bai J, Yin Z-D, Jiang H-Y, Guo Y-H, Ji DZD, Xu H-M, Li G, Mu GGC, Luo H-M, Wang J-L, Wang J, Ye X-M, Jin ZMY, Zhang W, Ning G-J, Wang H-Y, Li G-C, Yong J, Liang X-F, Liang G-D. 2011. Japanese Encephalitis, Tibet, China. Emerg Infect Dis 17:934– 936. doi:10.3201/eid1705.101417

Martins AS, Carvalho FA, Faustino AF, Martins IC, Santos NC. 2019. West Nile Virus Capsid Protein Interacts With Biologically Relevant Host Lipid Systems. Frontiers in Cellular and Infection Microbiology 9.

Mollentze N, Babayan SA, Streicker DG. n.d. Identifying and prioritizing potential human-infecting viruses from their genome sequences. PLOS BIOLOGY 25.

Ni X-B, Cui X-M, Liu J-Y, Ye R-Z, Wu Y-Q, Jiang J-F, Sun Y, Wang Q, Shum MH-H, Chang Q-C, Zhao L, Han X-H, Ma K, Shen S-J, Zhang M-Z, Guo W-B, Zhu J-G, Zhan L, Li L-J, Ding S-J, Zhu D-Y, Zhang J, Xia L-Y, Oong X-Y, Ruan X-D, Shao H-Z, Que T-C, Liu G-Y, Du C-H, Huang E-J, Wang X, Du L-F, Wang C-C, Shi W-Q, Pan Y-S, Zhou Y-H, Qu J-L, Ma J, Gong C-W, Chen Q-Q, Qin Q, Tick Genome and Microbiome Consortium (TIGMIC), Lam TT-Y, Jia N, Cao W-C. 2023. Metavirome of 31 tick species provides a compendium of 1,801 RNA virus genomes. Nat Microbiol 8:162–173. doi:10.1038/s41564-022-01275-w

Nouri S, Matsumura EE, Kuo Y-W, Falk BW. 2018. Insect-specific viruses: from discovery to potential translational applications. Current Opinion in Virology, Virus vector interactions • Special Section: Multicomponent viral systems 33:33–41. doi:10.1016/j.coviro.2018.07.006

Palacios G, Savji N, Travassos da Rosa A, Guzman H, Yu X, Desai A, Rosen GE, Hutchison S, Lipkin WI, Tesh R. 2013. Characterization of the Uukuniemi virus group (Phlebovirus: Bunyaviridae): evidence for seven distinct species. J Virol 87:3187– 3195. doi:10.1128/JVI.02719-12

Roth GA, Abate D, Abate KH, Abay SM, Abbafati C, Abbasi N, Abbastabar H, Abd-Allah F, Abdela J, Abdelalim A, Abdollahpour I, Abdulkader RS, Abebe HT, Abebe M, Abebe Z, Abejie AN, Abera SF, Abil OZ, Abraha HN, Abrham AR, Abu-Raddad LJ, Accrombessi MMK, Acharya D, Adamu AA, Adebayo OM, Adedoyin RA, Adekanmbi V, Adetokunboh OO, Adhena BM, Adib MG, Admasie A, Afshin A, Agarwal G, Agesa KM, Agrawal A, Agrawal S, Ahmadi A, Ahmadi M, Ahmed MB, Ahmed S, Aichour AN, Aichour I, Aichour MTE, Akbari ME, Akinyemi RO, Akseer N, Al-Aly Z, Al-Eyadhy A, Al-Raddadi RM, Alahdab F, Alam K, Alam T, Alebel A, Alene KA, Alijanzadeh M, Alizadeh-Navaei R, Aljunid SM, Alkerwi A, Alla F, Allebeck P, Alonso J, Altirkawi K, Alvis-Guzman N, Amare AT, Aminde LN, Amini E, Ammar W, Amoako YA, Anber NH, Andrei CL, Androudi S, Animut MD, Anjomshoa M, Ansari H, Ansha MG, Antonio CAT, Anwari P, Aremu O, Ärnlöv J, Arora A, Arora M, Artaman A, Aryal KK, Asayesh H, Asfaw ET, Ataro Z, Atique S, Atre SR, Ausloos M, Avokpaho EFGA, Awasthi A, Quintanilla BPA, Ayele Y, Ayer R, Azzopardi PS, Babazadeh A, Bacha U, Badali H, Badawi A, Bali AG, Ballesteros KE, Banach M, Banerjee K, Bannick MS, Banoub JAM, Barboza MA, Barker-Collo SL, Bärnighausen TW, Barquera S, Barrero LH, Bassat Q, Basu S, Baune BT, Baynes HW, Bazargan-Hejazi S, Bedi N, Beghi E, Behzadifar Masoud, Behzadifar Meysam, Béjot Y, Bekele BB, Belachew AB, Belay E, Belay YA, Bell ML, Bello AK, Bennett DA, Bensenor IM, Berman AE, Bernabe E, Bernstein RS, Bertolacci GJ, Beuran M, Beyranvand T, Bhalla A, Bhattarai S, Bhaumik S, Bhutta ZA, Biadgo B, Biehl MH, Bijani A, Bikbov B, Bilano V, Bililign N, Bin Sayeed MS, Bisanzio D, Biswas T, Blacker BF, Basara BB, Borschmann R, Bosetti C, Bozorgmehr K, Brady OJ, Brant LC, Brayne C, Brazinova A, Breitborde NJK, Brenner H, Briant PS, Britton G, Brugha T, Busse R, Butt ZA, Callender CSKH, Campos-Nonato IR, Campuzano Rincon JC, Cano J, Car M, Cárdenas R, Carreras G, Carrero JJ, Carter A, Carvalho F, Castañeda-Orjuela CA, Castillo Rivas J, Castle CD, Castro C, Castro F, Catalá-López F, Cerin E, Chaiah Y, Chang J-C, Charlson FJ, Chaturvedi P, Chiang PP-C, Chimed-Ochir O, Chisumpa VH, Chitheer A, Chowdhury R, Christensen H, Christopher DJ, Chung S-C, Cicuttini FM, Ciobanu LG, Cirillo M, Cohen AJ, Cooper LT, Cortesi PA, Cortinovis M, Cousin E, Cowie BC, Criqui MH, Cromwell EA, Crowe CS, Crump JA, Cunningham M, Daba AK, Dadi AF, Dandona L, Dandona R, Dang AK, Dargan PI, Daryani A, Das SK, Gupta RD, Neves JD, Dasa TT, Dash AP, Davis AC, Davis Weaver N, Davitoiu DV, Davletov K, De La Hoz FP, De Neve J-W, Degefa MG, Degenhardt L, Degfie TT, Deiparine S, Demoz GT, Demtsu BB, Denova-Gutiérrez E, Deribe K, Dervenis N, Des Jarlais DC, Dessie GA, Dey S, Dharmaratne SD, Dicker D, Dinberu MT, Ding EL, Dirac MA, Djalalinia S, Dokova K, Doku DT, Donnelly CA, Dorsey ER, Doshi PP, Douwes-Schultz D, Doyle KE, Driscoll TR, Dubey M, Dubljanin E, Duken EE, Duncan BB, Duraes AR, Ebrahimi H, Ebrahimpour S, Edessa D, Edvardsson D, Eggen AE, El Bcheraoui C, El Sayed Zaki M, El-Khatib Z, Elkout H, Ellingsen CL, Endres M, Endries AY, Er B, Erskine HE, Eshrati B, Eskandarieh S, Esmaeili R, Esteghamati A, Fakhar M, Fakhim H, Faramarzi M, Fareed M, Farhadi F, Farinha CSES, Faro A, Farvid MS, Farzadfar F, Farzaei MH, Feigin VL, Feigl AB, Fentahun N, Fereshtehnejad S-M, Fernandes E, Fernandes JC, Ferrari AJ, Feyissa GT, Filip I, Finegold S, Fischer F, Fitzmaurice C, Foigt NA, Foreman KJ, Fornari C, Frank TD, Fukumoto T, Fuller JE, Fullman N, Fürst T, Furtado JM, Futran ND, Gallus S, Garcia-Basteiro AL, Garcia-Gordillo MA, Gardner WM, Gebre AK, Gebrehiwot TT, Gebremedhin AT, Gebremichael B, Gebremichael TG, Gelano TF, Geleijnse JM, Genova-Maleras R, Geramo YCD, Gething PW, Gezae KE, Ghadami MR, Ghadimi R, Ghasemi Falavarjani K, Ghasemi-Kasman M, Ghimire M, Gibney KB, Gill PS, Gill TK, Gillum RF, Ginawi IA, Giroud M, Giussani G, Goenka S, Goldberg EM, Goli S, Gómez-Dantés H, Gona PN, Gopalani SV, Gorman TM, Goto A, Goulart AC, Gnedovskaya EV, Grada A, Grosso G, Gugnani HC, Guimaraes ALS, Guo Y, Gupta PC, Gupta Rahul, Gupta Rajeev, Gupta T, Gutiérrez RA, Gyawali B, Haagsma JA, Hafezi-Nejad N, Hagos TB, Hailegiyorgis TT, Hailu GB, Haj-Mirzaian Arvin, Haj-Mirzaian Arya, Hamadeh RR, Hamidi S, Handal AJ, Hankey GJ, Harb HL, Harikrishnan S, Haro JM, Hasan M, Hassankhani H, Hassen HY, Havmoeller R, Hay RJ, Hay SI, He Y, Hedayatizadeh-Omran A, Hegazy MI, Heibati B, Heidari M, Hendrie D, Henok A, Henry NJ, Herteliu C, Heydarpour F, Heydarpour P, Heydarpour S, Hibstu DT, Hoek HW, Hole MK, Homaie Rad E, Hoogar P, Hosgood HD, Hosseini SM, Hosseinzadeh M, Hostiuc M, Hostiuc S, Hotez PJ, Hoy DG, Hsiao T, Hu G, Huang JJ, Husseini A, Hussen MM, Hutfless S, Idrisov B, Ilesanmi OS, Iqbal U, Irvani SSN, Irvine CMS, Islam N, Islam SMS, Islami F, Jacobsen KH, Jahangiry L, Jahanmehr N, Jain SK, Jakovljevic M, Jalu MT, James SL, Javanbakht M, Jayatilleke AU, Jeemon P, Jenkins KJ, Jha RP, Jha V, Johnson CO, Johnson SC, Jonas JB, Joshi A, Jozwiak JJ, Jungari SB, Jürisson M, Kabir Z, Kadel R, Kahsay A, Kalani R, Karami M, Karami Matin B, Karch A, Karema C, Karimi-Sari H, Kasaeian A, Kassa DH, Kassa GM, Kassa TD, Kassebaum NJ, Katikireddi SV, Kaul A, Kazemi Z, Karyani AK, Kazi DS, Kefale AT, Keiyoro PN, Kemp GR, Kengne AP, Keren A, Kesavachandran CN, Khader YS, Khafaei B, Khafaie MA, Khajavi A, Khalid N, Khalil IA, Khan EA, Khan MS, Khan MA, Khang Y-H, Khater MM, Khoja AT, Khosravi A, Khosravi MH, Khubchandani J, Kiadaliri AA, Kibret GD, Kidanemariam ZT, Kiirithio DN, Kim D, Kim Y-E, Kim YJ, Kimokoti RW, Kinfu Y, Kisa A, Kissimova-Skarbek K, Kivimäki M, Knudsen AKS, Kocarnik JM, Kochhar S, Kokubo Y, Kolola T, Kopec JA, Koul PA, Koyanagi A, Kravchenko MA, Krishan K, Kuate Defo B, Kucuk Bicer B, Kumar GA, Kumar M, Kumar P, Kutz MJ, Kuzin I, Kyu HH, Lad DP, Lad SD, Lafranconi A, Lal DK, Lalloo R, Lallukka T, Lam JO, Lami FH, Lansingh VC, Lansky S, Larson HJ, Latifi A, Lau KM-M, Lazarus JV, Lebedev G, Lee PH, Leigh J, Leili M, Leshargie CT, Li S, Li Y, Liang J, Lim L-L, Lim SS, Limenih MA, Linn S, Liu S, Liu Y, Lodha R, Lonsdale C, Lopez AD, Lorkowski S, Lotufo PA, Lozano R, Lunevicius R, Ma S, Macarayan ERK, Mackay MT, MacLachlan JH, Maddison ER, Madotto F, Magdy Abd El Razek H, Magdy Abd El Razek M, Maghavani DP, Majdan M, Majdzadeh R, Majeed A, Malekzadeh R, Malta DC, Manda A-L, Mandarano-Filho LG, Manguerra H, Mansournia MA, Mapoma CC, Marami D, Maravilla JC, Marcenes W, Marczak L, Marks A, Marks GB, Martinez G, Martins-Melo FR, Martopullo I, März W, Marzan MB, Masci JR, Massenburg BB, Mathur MR, Mathur P, Matzopoulos R, Maulik PK, Mazidi M, McAlinden C, McGrath JJ, McKee M, McMahon BJ, Mehata S, Mehndiratta MM, Mehrotra R, Mehta KM, Mehta V, Mekonnen TC, Melese A, Melku M, Memiah PTN, Memish ZA, Mendoza W, Mengistu DT, Mengistu G, Mensah GA, Mereta ST, Meretoja A, Meretoja TJ, Mestrovic T, Mezgebe HB, Miazgowski B, Miazgowski T, Millear AI, Miller TR, Miller-Petrie MK, Mini GK, Mirabi P, Mirarefin M, Mirica A, Mirrakhimov EM, Misganaw AT, Mitiku H, Moazen B, Mohammad KA, Mohammadi M, Mohammadifard N, Mohammed MA, Mohammed S, Mohan V, Mokdad AH, Molokhia M, Monasta L, Moradi G, Moradi-Lakeh M, Moradinazar M, Moraga P, Morawska L, Moreno Velásquez I, Morgado-Da-Costa J, Morrison SD, Moschos MM, Mouodi S, Mousavi SM, Muchie KF, Mueller UO, Mukhopadhyay S, Muller K, Mumford JE, Musa J, Musa KI, Mustafa G, Muthupandian S, Nachega JB, Nagel G, Naheed A, Nahvijou A, Naik G, Nair S, Najafi F, Naldi L, Nam HS, Nangia V, Nansseu JR, Nascimento BR, Natarajan G, Neamati N, Negoi I, Negoi RI, Neupane S, Newton CRJ, Ngalesoni FN, Ngunjiri JW, Nguyen AQ, Nguyen G, Nguyen Ha Thu, Nguyen Huong Thanh, Nguyen LH, Nguyen M, Nguyen TH, Nichols E, Ningrum DNA, Nirayo YL, Nixon MR, Nolutshungu N, Nomura S, Norheim OF, Noroozi M, Norrving B, Noubiap JJ, Nouri HR, Nourollahpour Shiadeh M, Nowroozi MR, Nyasulu PS, Odell CM, Ofori-Asenso R, Ogbo FA, Oh I-H, Oladimeji O, Olagunju AT, Olivares PR, Olsen HE, Olusanya BO, Olusanya JO, Ong KL, Ong SKS, Oren E, Orpana HM, Ortiz A, Ortiz JR, Otstavnov SS, Øverland S, Owolabi MO, Özdemir R, P A M, Pacella R, Pakhale S, Pakhare AP, Pakpour AH, Pana A, Panda-Jonas S, Pandian JD, Parisi A, Park E-K, Parry CDH, Parsian H, Patel S, Pati S, Patton GC, Paturi VR, Paulson KR, Pereira A, Pereira DM, Perico N, Pesudovs K, Petzold M, Phillips MR, Piel FB, Pigott DM, Pillay JD, Pirsaheb M, Pishgar F, Polinder S, Postma MJ, Pourshams A, Poustchi H, Pujar A, Prakash S, Prasad N, Purcell CA, Qorbani M, Quintana H, Quistberg DA, Rade KW, Radfar A, Rafay A, Rafiei A, Rahim F, Rahimi K, Rahimi-Movaghar A, Rahman M, Rahman MHU, Rahman MA, Rai RK, Rajsic S, Ram U, Ranabhat CL, Ranjan P, Rao PC, Rawaf DL, Rawaf S, Razo-García C, Reddy KS, Reiner RC, Reitsma MB, Remuzzi G, Renzaho AMN, Resnikoff S, Rezaei S, Rezaeian S, Rezai MS, Riahi SM, Ribeiro ALP, Rios-Blancas MJ, Roba KT, Roberts NLS, Robinson SR, Roever L, Ronfani L, Roshandel G, Rostami A, Rothenbacher D, Roy A, Rubagotti E, Sachdev PS, Saddik B, Sadeghi E, Safari H, Safdarian M, Safi S, Safiri S, Sagar R, Sahebkar A, Sahraian MA, Salam N, Salama JS, Salamati P, Saldanha RDF, Saleem Z, Salimi Y, Salvi SS, Salz I, Sambala EZ, Samy AM, Sanabria J, Sanchez-Niño MD, Santomauro DF, Santos IS, Santos JV, Milicevic MMS, Sao Jose BP, Sarker AR, Sarmiento-Suárez R, Sarrafzadegan N, Sartorius B, Sarvi S, Sathian B, Satpathy M, Sawant AR, Sawhney M, Saxena S, Sayyah M, Schaeffner E, Schmidt MI, Schneider IJC, Schöttker B, Schutte AE, Schwebel DC, Schwendicke F, Scott JG, Sekerija M, Sepanlou SG, Serván-Mori E, Seyedmousavi S, Shabaninejad H, Shackelford KA, Shafieesabet A, Shahbazi M, Shaheen AA, Shaikh MA, Shams-Beyranvand M, Shamsi M, Shamsizadeh M, Sharafi K, Sharif M, Sharif-Alhoseini M, Sharma R, She J, Sheikh A, Shi P, Shiferaw MS, Shigematsu M, Shiri R, Shirkoohi R, Shiue I, Shokraneh F, Shrime MG, Si S, Siabani S, Siddiqi TJ, Sigfusdottir ID, Sigurvinsdottir R, Silberberg DH, Silva DAS, Silva JP, Silva NTD, Silveira DGA, Singh JA, Singh NP, Singh PK, Singh V, Sinha DN, Sliwa K, Smith M, Sobaih BH, Sobhani S, Sobngwi E, Soneji SS, Soofi M, Sorensen RJD, Soriano JB, Soyiri IN, Sposato LA, Sreeramareddy CT, Srinivasan V, Stanaway JD, Starodubov VI, Stathopoulou V, Stein DJ, Steiner C, Stewart LG, Stokes MA, Subart ML, Sudaryanto A, Sufiyan MB, Sur PJ, Sutradhar I, Sykes BL, Sylaja PN, Sylte DO, Szoeke CEI, Tabarés-Seisdedos R, Tabuchi T, Tadakamadla SK, Takahashi K, Tandon N, Tassew SG, Taveira N, Tehrani-Banihashemi A, Tekalign TG, Tekle MG, Temsah M-H, Temsah O, Terkawi AS, Teshale MY, Tessema B, Tessema GA, Thankappan KR, Thirunavukkarasu S, Thomas N, Thrift AG, Thurston GD, Tilahun B, To QG, Tobe-Gai R, Tonelli M, Topor-Madry R, Torre AE, Tortajada-Girbés M, Touvier M, Tovani-Palone MR, Tran BX, Tran KB, Tripathi S, Troeger CE, Truelsen TC, Truong NT, Tsadik AG, Tsoi D, Tudor Car L, Tuzcu EM, Tyrovolas S, Ukwaja KN, Ullah I, Undurraga EA, Updike RL, Usman MS, Uthman OA, Uzun SB, Vaduganathan M, Vaezi A, Vaidya G, Valdez PR, Varavikova E, Vasankari TJ, Venketasubramanian N, Villafaina S, Violante FS, Vladimirov SK, Vlassov V, Vollset SE, Vos T, Wagner GR, Wagnew FS, Waheed Y, Wallin MT, Walson JL, Wang Y, Wang Y-P, Wassie MM, Weiderpass E, Weintraub RG, Weldegebreal F, Weldegwergs KG, Werdecker A, Werkneh AA, West TE, Westerman R, Whiteford HA, Widecka J, Wilner LB, Wilson S, Winkler AS, Wiysonge CS, Wolfe CDA, Wu S, Wu Y-C, Wyper GMA, Xavier D, Xu G, Yadgir S, Yadollahpour A, Yahyazadeh Jabbari SH, Yakob B, Yan LL, Yano Y, Yaseri M, Yasin YJ, Yentür GK, Yeshaneh A, Yimer EM, Yip P, Yirsaw BD, Yisma E, Yonemoto N, Yonga G, Yoon S-J, Yotebieng M, Younis MZ, Yousefifard M, Yu C, Zadnik V, Zaidi Z, Zaman SB, Zamani M, Zare Z, Zeleke AJ, Zenebe ZM, Zhang AL, Zhang K, Zhou M, Zodpey S, Zuhlke LJ, Naghavi M, Murray CJL. 2018. Global, regional, and national age-sex-specific mortality for 282 causes of death in 195 countries and territories, 1980–2017: a systematic analysis for the Global Burden of Disease Study 2017. The Lancet 392:1736–1788. doi:10.1016/S0140-6736(18)32203-7

Sato Y, Mekata H, Sudaryatma PE, Kirino Y, Yamamoto S, Ando S, Sugimoto T, Okabayashi T. 2021. Isolation of Severe Fever with Thrombocytopenia Syndrome Virus from Various Tick Species in Area with Human Severe Fever with Thrombocytopenia Syndrome Cases. Vector-Borne and Zoonotic Diseases 21:378–384. doi:10.1089/vbz.2020.2720

Sen R, Nayak L, De RK. 2016. A review on host–pathogen interactions: classification and prediction. Eur J Clin Microbiol Infect Dis 35:1581–1599. doi:10.1007/s10096-016-2716-7

Stephenson EB, Murphy AK, Jansen CC, Peel AJ, McCallum H. 2019. Interpreting mosquito feeding patterns in Australia through an ecological lens: an analysis of blood meal studies. Parasites & Vectors 12:156. doi:10.1186/s13071-019-3405-z

Tabachnick WJ. 2016. Climate Change and the Arboviruses: Lessons from the Evolution of the Dengue and Yellow Fever Viruses. Annual Review of Virology 3:125–145. doi:10.1146/annurev-virology-110615-035630

Touray M, Bakirci S, Ulug D, Gulsen SH, Cimen H, Yavasoglu SI, Simsek FM, Ertabaklar H, Ozbel Y, Hazir S. 2023. Arthropod vectors of disease agents: Their role in public and veterinary health in Turkiye and their control measures. Acta Tropica 243:106893. doi:10.1016/j.actatropica.2023.106893

Turner EA, Christofferson RC. 2024. Exploring the transmission modalities of Bunyamwera virus. Exp Biol Med (Maywood) 249:10114. doi:10.3389/ebm.2024.10114

Vector-borne diseases. n.d. https://www.who.int/news-room/fact-sheets/detail/vector-borne-diseases

Viglietta M, Bellone R, Blisnick AA, Failloux A-B. 2021. Vector Specificity of Arbovirus Transmission. Frontiers in Microbiology 12.

Vogels CBF, Rückert C, Cavany SM, Perkins TA, Ebel GD, Grubaugh ND. 2019. Arbovirus coinfection and co-transmission: A neglected public health concern? PLOS Biology 17:e3000130. doi:10.1371/journal.pbio.3000130

Weaver SC, Charlier C, Vasilakis N, Lecuit M. 2018. Zika, Chikungunya, and Other Emerging Vector-Borne Viral Diseases. Annu Rev Med 69:395–408. doi:10.1146/annurev-med-050715-105122

Weissenböck H, Hubálek Z, Bakonyi T, Nowotny N. 2010. Zoonotic mosquito-borne flaviviruses: Worldwide presence of agents with proven pathogenicity and potential candidates of future emerging diseases. Veterinary Microbiology 140:271–280. doi:10.1016/j.vetmic.2009.08.025

Wu Z, Zhang M, Zhang Y, Lu K, Zhu W, Feng S, Qi J, Niu G. 2023. Jingmen tick virus: an emerging arbovirus with a global threat. mSphere 8:e00281–23. doi:10.1128/msphere.00281-23

Xia H, Liu R, Zhao L, Sun X, Zheng Z, Atoni E, Hu X, Zhang B, Zhang G, Yuan Z. 2020. Characterization of Ebinur Lake Virus and Its Human Seroprevalence at the China– Kazakhstan Border. Frontiers in Microbiology 10.

Yang C, Wang F, Huang D, Ma H, Zhao L, Zhang G, Li H, Han Q, Bente D, Salazar FV, Yuan Z, Xia H. 2022. Vector competence and immune response of Aedes aegypti for Ebinur Lake virus, a newly classified mosquito-borne orthobunyavirus. PLoS Negl Trop Dis 16:e0010642. doi:10.1371/journal.pntd.0010642

Yang X, Qin S, Liu X, Zhang N, Chen J, Jin M, Liu F, Wang Y, Guo J, Shi H, Wang C, Chen Y. 2023. Meta-Viromic Sequencing Reveals Virome Characteristics of Mosquitoes and Culicoides on Zhoushan Island, China. Microbiol Spectr e02688–22. doi:10.1128/spectrum.02688-22

Zaid A, Burt FJ, Liu X, Poo YS, Zandi K, Suhrbier A, Weaver SC, Texeira MM, Mahalingam S. 2021. Arthritogenic alphaviruses: epidemiological and clinical perspective on emerging arboviruses. The Lancet Infectious Diseases 21:e123–e133. doi:10.1016/S1473-3099(20)30491-6

Zhang Zheng, Cai Z, Tan Z, Lu C, Jiang T, Zhang G, Peng Y. 2019. Rapid identification of human -infecting viruses. Transbound Emerg Dis 66:2517 – 2522. doi:10.1111/tbed.13314

Zhang Zhenzhen, Rong L, Li Y-P. 2019. Flaviviridae Viruses and Oxidative Stress: Implications for Viral Pathogenesis. Oxid Med Cell Longev 2019:1409582. doi:10.1155/2019/1409582

Zhao L, Luo H, Huang D, Yu P, Dong Q, Mwaliko C, Atoni E, Nyaruaba R, Yuan J, Zhang G, Bente D, Yuan Z, Xia H. 2020. Pathogenesis and Immune Response of Ebinur Lake Virus: A Newly Identified Orthobunyavirus That Exhibited Strong Virulence in Mice. Front Microbiol 11:625661. doi:10.3389/fmicb.2020.625661

Zhao L, Yu P, Shi C, Jia L, Evans A, Wang X, Wu Q, Xiong G, Ming Z, Salazar F, Agwanda B, Bente D, Wang F, Liu D, Yuan Z, Xia H. 2022. Global mosquito virome profiling and mosquito spatial diffusion pathways revealed by marker-viruses (preprint). Microbiology. doi:10.1101/2022.09.24.509300

